# Frequency-Domain Evaluation and Estimation of Semi-Markov Disease Natural History Models

**DOI:** 10.64898/2026.10.23.26366343

**Authors:** Felipe Subiabre, Jeremy D. Goldhaber-Fiebert, Fernando Alarid-Escudero

## Abstract

**Introduction:** Natural history models must often be estimated from data that do not directly capture disease transitions. Semi-Markov processes let the risk of progression depend on the time spent in a health state, which Markov models need extra states to represent. Their limitation is computational: discretizing time introduces approximation error, and simulating individuals introduces Monte Carlo error and a non-differentiable calibration objective.

**Methods:** We show that the model-implied prevalence, incidence and cause-specific mortality of a continuous-time semi-Markov model satisfy convolution equations that become a small linear system when Laplace-transformed. Solving it on a grid of complex frequencies and inverting with an accelerated Fourier series returns the targets without a time grid or Monte Carlo error, and yields exact first- and second-order derivatives for one additional linear solve each. These support Newton-type maximum likelihood estimation, gradient-informed Bayesian sampling, and local identifiability diagnostics. We apply our open-source implementation to a urinary bladder cancer model calibrated to SEER incidence and mortality by age and stage.

**Results:** The method reproduced an independent implementation to within its accuracy (maximum relative difference 8 × 10^−6^) and evaluated all targets, their gradient, and their Hessian in 1.3, 2.5, and 9.1 milliseconds; a one-year-cycle cohort approximation deviated by up to 42% and a two-million-person microsimulation by up to 811%. Maximum likelihood and Bayesian estimates agreed, and second-order information showed that the aggregate targets constrain where individuals go from preclinical states far better than how long they stay. Early-stage incidence and mortality showed systematic misfit across age.

**Conclusions:** The framework supplies existing calibration algorithms with fast, exact model evaluations and derivatives for models whose transition hazards depend on time in state rather than on age.

**Highlights:**

- The age-specific prevalence, incidence, and cause-specific mortality implied by a continuous-time semi-Markov model can be computed in the frequency domain without time discretization and without Monte Carlo error.
- The same computation returns exact gradients and Hessians of the calibration objective, which enables Newton-type optimization, gradient-informed Bayesian calibration, and local identifiability diagnostics that derivative-free calibration cannot provide.
- The framework supplies model evaluations and derivatives to existing calibration algorithms rather than replacing them, as we demonstrate using a urinary bladder cancer model fitted to aggregate SEER data.
- Open-source software implements the framework, including competing risks and age-specific background mortality rates.

## 1 Introduction

Natural history models describe how a disease progresses through health states in the absence of intervention and form the foundation for much of the modeling in medical decision-making [1]. Their central difficulty is that the natural history is largely unobserved: registries record diagnosis, treatment, and death, not the onset of disease or its progression through preclinical stages. The parameters that govern progression must therefore be inferred indirectly, from population quantities such as age- and stage-specific incidence and mortality, which are often available only in aggregate [2] [3].

The model structure determines both what can be inferred and the computational cost. Cohort Markov models are computationally inexpensive but memoryless: the risk of leaving a state cannot depend on how long a patient has been in it unless additional tunnel states are added [4]. Individual-level microsimulation removes this restriction but introduces Monte Carlo error into every output, especially for rare targets, and results in a stochastic calibration objective without usable derivatives [5]. Semi-Markov models are a middle ground [6, 7]. Their transition hazards depend on time spent in states, allowing parameters to model various dwell-time patterns while remaining deterministic and cohort-level. They are used in health economics [8] and multi-state survival analysis [2, 3], where the estimation is based on individual time-to-event data, similar to Bayesian methods for Markov renewal processes [9]. Related theory has been applied to models of coronary patients [10].

What is missing is not a method for calibrating models to aggregate data. Bayesian calibration, approximate Bayesian computation, and incremental mixture importance sampling accept aggregate targets, with their likelihood or distance functions chosen to match the data [11, 12, 13]. Each method, however, treats the model as a black box, incurs a computational cost at every evaluation, and cannot use derivatives. For continuous-time semi-Markov models, an exact, fast, and differentiable solution has been lacking.

In this manuscript, we propose such a solution. The model-implied targets satisfy convolution equations whose Laplace transform forms a linear system with one equation per state. Thus, all age-specific targets follow from a small linear solve per frequency and an accelerated Fourier inversion, with no time grid and no simulation. Differentiating the same system yields exact gradients and Hessians of the calibration objective. We illustrate the framework with a urinary bladder cancer model calibrated to SEER data, verify it against an independent implementation, and provide open-source software.

## 2 Methods

### 2.1 Overview of the approach

Before formal development, we describe what the framework computes and why it operates in the frequency domain. Figure 1 summarizes the sequence.

**Figure 1:**
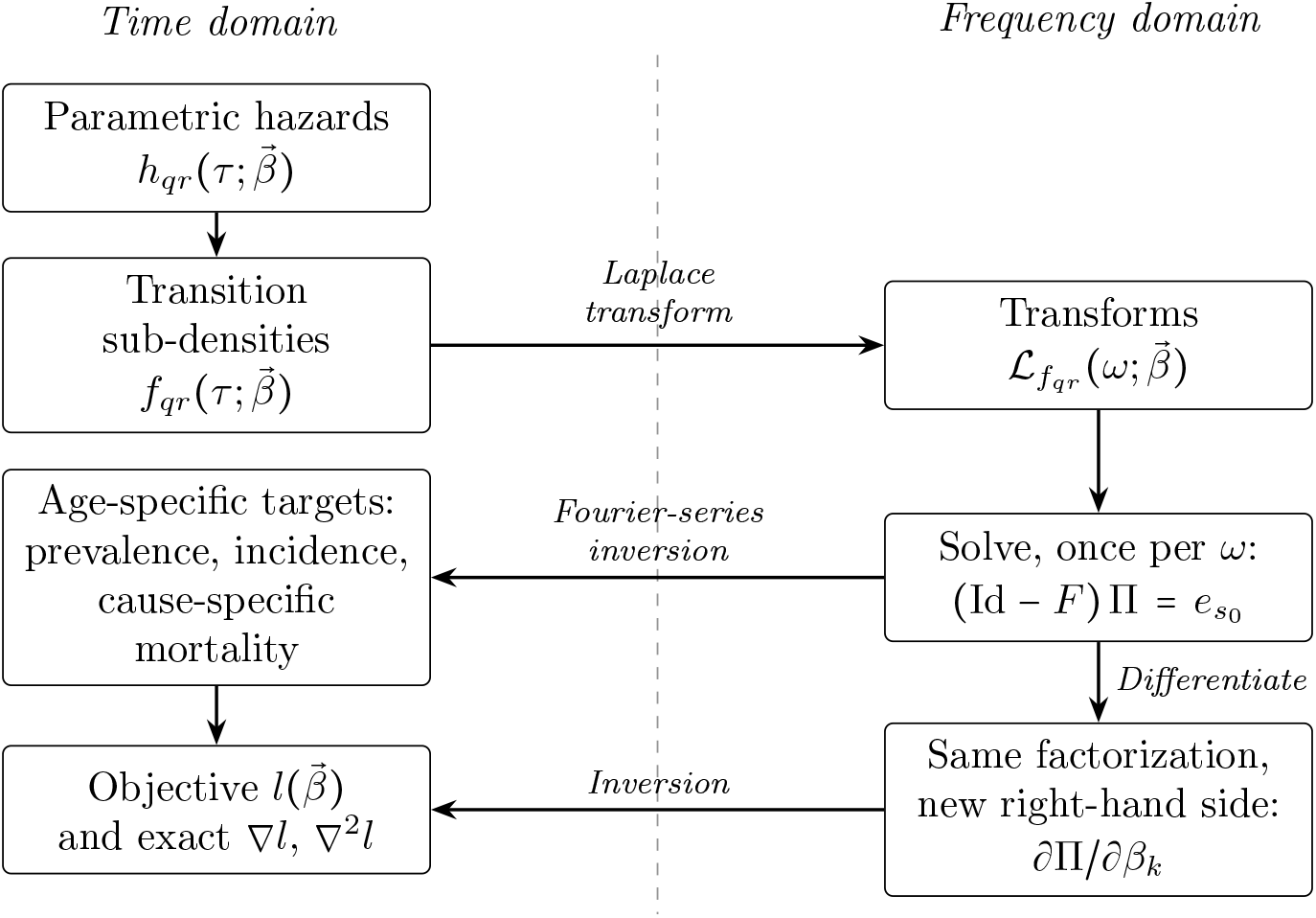
Calculation of model-implied targets and derivatives involves converting parametric transition hazards into transition sub-densities, which are then Laplace-transformed. We solve a linear system of dimension ∣*S*∣ with coefficient matrix *F* (*ω*) representing the transformed sub-densities at several complex frequencies to obtain the transformed arrival densities Π (*ω*) . Derivatives with respect to parameters are computed by solving the same system with a different right-hand side, reusing the factorization. An accelerated Fourier series then provides the age-specific targets, enabling the assembly of the calibration objective, its exact gradient, and Hessian. This process involves no time grid and requires no simulation of quantities.

The modeler specifies a set of health states and, for each allowed transition, a parametric hazard that depends on the time spent in the origin state. From these inputs, the framework computes the model-predicted quantities that match aggregate data, such as the agespecific proportion of the cohort in each state, the arrival rate in each state, and the death rate by cause, along with their derivatives with respect to the parameters (Figure 1). Sections 2.2.1 to 2.4 develop the evaluation, Section 2.5 builds the calibration targets, Sections 2.6 to 2.8 cover derivatives, identifiability, and estimation, and Section 2.9 relates the framework to existing calibration approaches.

### 2.2 Model dynamics and target functions

#### 2.2.1 Setup

The disease model is a semi-Markov continuous-time process *X*_*t t* ≥0_, a class that generalizes both discrete-time Markov chains and continuous-time, time-homogeneous Markov processes. It evolves over a finite discrete set of states *S*, from an initial state *S*_0_ ∈ *S*, and a subset of absorbing *death states D* ⊆ *S*.

The *holding time* (also called the *sojourn time* or *dwell time*) of a state *q* is the non-negative random variable *ϕ*_*q*_ that measures the time spent in *q* before transitioning to the next state in the process. Each possible transition between states *q* and *r* in *S* is described by a nonnegative hazard rate *h*_*qr*_(*τ*) , where *τ* ≥ 0 measures the *local time*, that is, the time spent in the current state *q* since the last transition to it. The value *h*_*qr*_ (*τ*) is the instantaneous rate of transitioning from state *q* to state *r* at local time *τ* , given that no transition has occurred by then

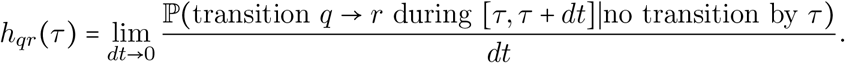

Therefore, two clocks run at once: *global time t*, the time since the start of the process (attained age in our application, with *t* = 0 at birth), and local time *τ* , which resets to zero at every transition.

Given the hazard rate functions *h*_*qr*_, the cumulative hazard functions *H*_*q*_ and marginal transition functions *f*_*qr*_ are given by [14, 15]:

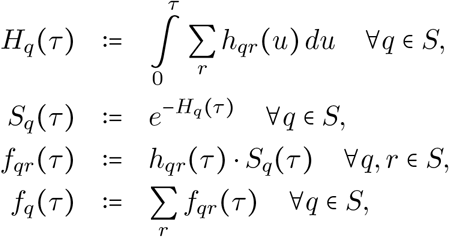

where the sums are understood to be taken over states connected from *q*. With these definitions, one can verify that *H*_*q*_ (∞) = ∞ implies 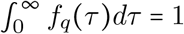. Unlike hazards, these functions are unconditional, where *f*_*q*_ is the density of the holding time *ϕ*_*q*_, whereas each *f*_*qr*_ is a *sub-density*, a density in the joint space of exit time and destination whose integral 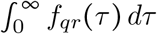 is the probability that the exit from *q* is to *r* [16, 17]. Only their sum *f*_*q*_ integrates to one.

For our present analysis, we consider parametric hazards of the form 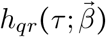, where the functional form of *h*_*qr*_ is known, 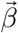 is a finite-dimensional real vector of unknown model parameters, the object of estimation, and *h*_*qr*_ is smooth on its domain. In practice, *h*_*qr*_ is chosen as the hazard rate function corresponding to a time-to-event distribution (e.g., exponential, log-normal, Weibull, Gompertz, etc.). Given the formulas above, the chosen parametric hazard rates translate into corresponding parametric transition subdensities. In the bladder cancer model of Section 3, for example, 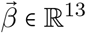 collects the shape and scale of a Weibull onset hazard and eleven constant hazard rates. It contains no individual-level covariates, and age is the time axis of the model.

#### 2.2.2 Notation

The indices *q, r, s* represent states in the semi-Markov process. We use *q* and *r* to represent states that are connected through a transition *q* → *r*. The indices *k, l* represent specific components of the model parameters. The function 1_*A*_ is the indicator function of the event *A*, equal to one if the event is true and 0 otherwise. Finally, for a given measurable function 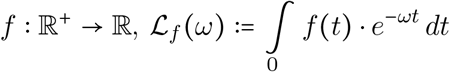 denotes its Laplace transform evaluated at a complex frequency *ω* ∈ C . The symbol · always denotes a product of scalars; the only vector and matrix objects are 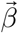, the vector Π *ω* and the matrix *F ω* , which appear onlyin matrix notation. Table 1 condenses the main notation used throughout the manuscript.

**Table 1:** Notation summary.

| Notation | Description |
| --- | --- |
| $t$ | Global time, time since start of the process (e.g., attained age) |
| $\tau$ | Local time, time since arrival at the current state |
| $X_t$ | State occupied at global time $t$ |
| $S, D, s_0$ | State space, absorbing death states, initial state |
| $q, r, s$ | Model states |
| $\beta; k, l$ | Vector of model parameters; indices of its components |
| $h_{qr}, H_q$ | Hazard of transition $q \rightarrow r$; cumulative hazard of leaving $q$ |
| $\phi_q, S_q$ | Holding time of state $q$; its survival function |
| $f_q, f_{qr}$ | Density of $\phi_q$; sub-density of leaving $q$ for $r$ |
| $\pi_s$ | Arrival density of state $s$ (Definition 1) |
| $p_s, g_s$ | Occupancy $\mathbb{P}(X_t = s)$; expected occupation time up to $t$ |
| $\mathcal{L}_f(\omega)$ | Laplace transform of $f$ evaluated at complex frequency $\omega$ |
| $\Pi_s, F$ | Transform of $\pi_s$; matrix with entries $F_{sq} = \mathcal{L}_{f_{qs}}$ |
| $\mathcal{S}, S_0, \lambda_0$ | Model survival; background survival and hazard |
| $\alpha$ | Continuous discount rate, $\alpha = \log(1 + r)$ for an annual rate $r$ |
| $\mathbb{1}_A$ | Indicator function of the event $A$ |
| $l(\vec{\beta})$ | Objective function |

### 2.3 Expressing the semi-Markov dynamics in frequency space

Three results underlie the computation. The first expresses the dynamics as a system of convolution equations for the rates at which individuals arrive in each state; the second solves that system in the frequency domain; the third recovers state occupancy and discounted occupation times.

Proofs are in the Supplementary Material, and [6] (Sections 3 and 4) gives a general treatment of these renewal equations.

We start by defining the two objects in which the dynamics are expressed.

#### Definition 1.

For a state *s* ∈ *S*, let *N*_*s*_ (*t*) count the arrivals of the process in during [0, *t*], with the initial placement in *s*_0_ at *t* = 0 counted as an arrival. The *arrival density* of *s* is

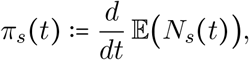

so that *π*_*s*_ (*t*) *dt* is the expected number of arrivals in *s* during [*t, t* + *dt*), and 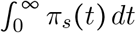 is the expected number of visits to *s*. When *s* can be visited at most once (as in the progressive model of Section 3), *π*_*s*_ is the sub-density of the age at arrival in *s*, with total mass equal to the probability of ever reaching *s*. The initial state contributes to the atom *δ*_0_, where the Dirac delta distribution at *t* = 0 equals to 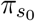. The *occupation time* of *s* up to global time *t* is

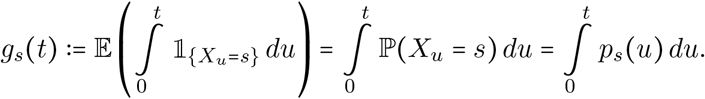

The arrival density is easily confused with two other quantities. It is not the distribution of the time *spent* in *s* during a visit, which is the holding time *ϕ*_*s*_, nor the proportion of the cohort *occupying s*, which is *p*_*s*_ ≔ P (*X*_*t*_ = *s*). It is the flow of individuals into *s*, and divided by the surviving population (Section 2.5), it is the incidence of state *s*.

#### Proposition 1.

The arrival densities (*π*_*s*_) _*s*∈ *S*_ of the process started in *s*_0_ at *t* = 0 are the unique solution of the system of convolution equations

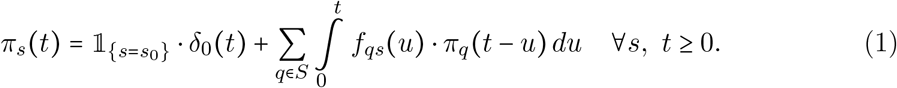

For each state *s* ∈ *S*, the occupation time *g*_*s*_ is the convolution of *π*_*s*_(*t*) and the function *d*_*s*_(*t*) ≔ E(min(*ϕ*_*s*_, *t*)).

*Proof*. See the Supplementary Material.

The system (1) is the Markov renewal equation of the process [6], and it reads as a statement about individual histories. An arrival in state *s* at age *t* is either the placement of the cohort in *s*_0_ at age zero (the Dirac term) or a transition from some state *q*. In the latter case, the individual arrived in at an earlier age *t* − *u*, at a rate *π*_*q*_(*t* − *u*), and left *q* for exactly after units of local time, with sub-density . Because the local clock resets on arrival in *q*, the two factors multiply, integrating over the unobserved age *t* − *u* and summing over *q* gives (1), which is the law of total probability applied to the last transition before *t*.

Two consequences are worth stating. First, (1) is not a hypothesis on the *π*_*s*_ (*u*): given the sub-densities *f*_*qs*_, the system has a unique solution when *S* is finite (Corollary 3.16 of [6]), so there is no admissibility condition to check. Second, an individual arriving in *s* at age *u* spends an expected time *d*_*s*_(*t* − *u*) in *s* before the horizon *t*; integrating against arrivals gives *g*_*s*_ = *π*_*s*_ ∗ *d*_*s*_.

Taking Laplace transforms in (1) turns each convolution into a product and the whole system into a linear one. Writing 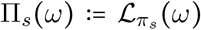 and collecting the transformed sub-densities into the matrix *F* (*ω*) with entries 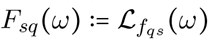, we obtain the following.

#### Corollary 2.

For any *ω* ∈ ℂ at which the transforms 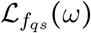 exist and Id − *F* (*ω*) is invertible, the vector Π(*ω*) = (Π_*s*_(*ω*))_*s*∈*S*_ is the unique solution of

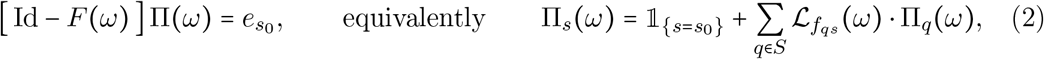

where 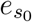 is the canonical vector of the initial state. For any *s* ∈ *S*,

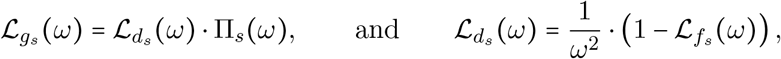

for any *ω* ∈ C at which the quantities involved exist uniquely.

*Proof*. The first and second claims follow from taking the Laplace transforms in Proposition 1, identifying 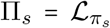 and using the convolution property; uniqueness holds wherever is invertible, which is the case on a right half-plane because Id − *F* (*ω*) as Re (*ω*) → ∞ . For a proof of the third claim, see the Supplementary Material.

Corollary 2 is where the computational gain is realized. Π_*s*_ (*ω*) has no epidemiological interpretation of its own: it is the Laplace transform of the arrival density of state *s*, a reencoding of the incidence flow that is far cheaper to compute. The time-domain system (1), whose direct solution requires nested integration along every path through the state space, becomes a linear system of dimension ∣*S*∣ at each frequency. One matrix factorization per frequency delivers the transform of every target; the age resolution is chosen afterward, in the inversion of Section 2.4, rather than by a time grid; and the parameter derivatives solve the same system with a different right-hand side (Section 2.6).

The third result recovers the occupancy of each state from its arrivals and identifies one case that requires no inversion: discounted lifetime occupation times, obtained by solving the system in Corollary 2 at the single real frequency *ω* = *α* , where *α* > 0 is the continuous discount rate (*α* = log (1 + *r*) for an annual rate *r*).

#### Corollary 3.

The population profile functions are the convolutions of each state’s arrival density with the survival function of its holding time:

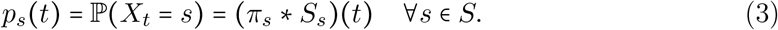

Moreover, for any *s* ∈ *S* and any *α* > 0 at which the transforms exist,

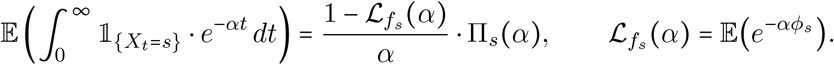

*Proof*. See the Supplementary Material.

This is the discounting used in health economic evaluation [18, 19, 20], with an annual discount rate , a value accruing years ahead is weighted by (1 + *r*)^−*t*^ = *e*^−*αt*^ with *α* = ln (1 + *r*), so that *r* = 5% gives *α* ≈ 0.0488, and *α* → 0^+^ recovers the undiscounted case. Discounted life years spent in each state, the building block of discounted, quality-adjusted life years, therefore, follow from Corollary 3 with no further computation.

*Example* 1 (A three-state progressive model in closed form). Let = *H*{, *P, C, D*_*c*_} with *s* = *H*_0_: healthy, preclinical, clinically detected, and death from disease. Suppose onset *H* → *P* has constant hazard , progression to detection *P* → *C* has constant hazard , and death before detection *P* → *D*_*c*_ competes with it at constant hazard . The holding time *ϕ*_*P*_ is then exponential with rate *b + m* and the sub-densities are

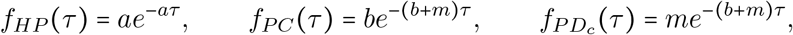

with transforms 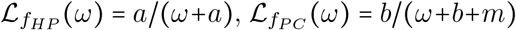 and 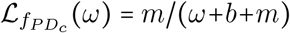. The linear system of Corollary 2 is triangular here and is solved by substitution:

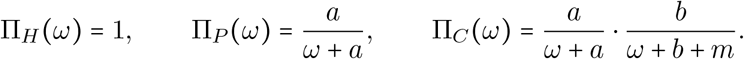

Inverting term by term,

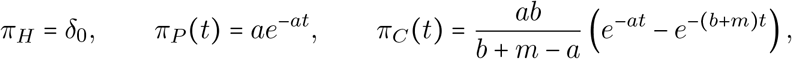

so the arrival density of *P* is the density of an exponential age at onset, as it must be, and the arrival density of *C* is a hypoexponential sub-density with total mass

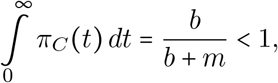

the proportion of the cohort ever detected rather than dying undetected first, *π*_*C*_, integrates to the lifetime risk of detection, not to one, which is the sub-density property of Section 2.2.1. The occupancy of the preclinical state follows from (3),

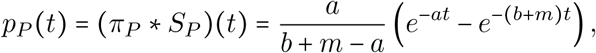

and the preclinical prevalence and clinical incidence targets of Section 2.5 are ratios of these expressions.

The example generalizes directly. If the onset hazard is Weibull, *h*_*HP*_ (*τ*) = (*k* / *λ*) (*τ* / *λ*)^*k* −1^, as in Section 3, every displayed equation above still holds. The only difference is that 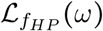 loses its closed form and is evaluated by one-dimensional quadrature per frequency, after which the linear algebra and the inversion of Section 2.4 are unchanged.

Algorithm 1 states the general computation that Example 1 carries out by hand. Steps 1 to 5 evaluate the objective; step 6 adds its exact derivatives (Section 2.6).

#### Algorithm 1

(Evaluation of model targets, objective and derivatives). *Input:* hazards 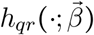 and the parameter value 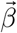; initial state *s*_0_; ages *t*_1_ < … < *t*_*n*_ at which targets are required; inversion parameters *A*, ℓ and number of terms *K*; observed targets and their standard errors.

1. Build the frequency grid *ω*_*j*_ = *A /* (2ℓ*t*_max_) + i *jπ /* (ℓ*t*_max_) , *j* = 0, …, *K*, where 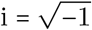 and *t*_max_ is the largest age at which a transform is inverted.
2. For each *ω*_*j*_: evaluate 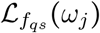 for every transition, in closed form where available and otherwise by one-dimensional quadrature; assemble *F* (*ω*_*j*_); factorize Id − *F* (*ω*_*j*_) once and and solve (2) for Π(*ω*_*j*_).
3. Form the transform of each required target by multiplication: 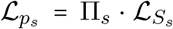 for occupancy, Π_*s*_/*ω* for cumulative arrivals, 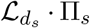 for occupation time, and 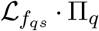 for the flow into a death state *s* from each origin *q*.
4. Invert each transform by the accelerated Fourier series of Section 2.4: for interval targets, invert the cumulative transforms Π_*s*_ (*ω*) / *ω* at the ages that bound each interval and at the life table’s half-year steps; for pointwise targets, invert at the required ages.
5. Assemble prevalence, incidence and cause-specific mortality as in Section 2.5 and evaluate 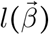 as in Section 2.5.5.
6. For derivatives: differentiate (2) with respect to *β*_*k*_ and reuse the factorization from step 2, one additional back-substitution per parameter for ∇*l* and one per parameter pair for ∇^2^*l*; then repeat steps 3 to 5 on the differentiated transforms.

*Cost: O* (*K* (∣*S*∣ ^3^ + *P* ^2^ ∣*S*∣ ^2^)) arithmetic operations plus *O* (*K*· {∣*q* → *r*∣ · *P* ^2^) quadratures for steps 2 and 3, independent of the number of ages *n* and of any time-step choice; the inversion in step 4 adds a cost linear in the number of time points at which the series is evaluated. Discounted lifetime quantities (Corollary 3) need only step 2 at the single frequency *ω* = *α*.

### 2.4 Recovering the dynamics in the time domain

To return to the time domain, we use the Fourier-series method for Laplace transforms of [21] (Section 1.2.2). For a function *ψ* on [0, *t*_max_] with transform ℒ_*ψ*_,

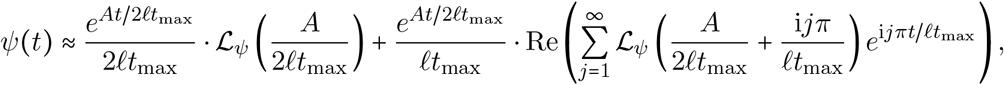

with the standard values *A* = 30 and ℓ = 5; the frequencies in the sum are the grid *ω*_*j*_ of Algorithm 1.

The series converges slowly, and [21] accelerate it with the Euler summation, which is effective because the functions being recovered are smooth. Instead, we adapt the Padé-type acceleration of [22], which converges exponentially faster.

For the model of Section 3, 120 series terms recover the arrival densities to a relative precision of about 2 × 10 ^−8^ at ages 35 to 80, away from *t* = 0, where the Gibbs phenomenon dominates (Supplementary Figure S1). Interval targets are obtained by inverting the cumulative transforms Π_*s*_ (*ω*) / *ω* directly at the ages that bound each interval, and at the half-year points where the life table changes value (Supplementary Section 6.1), so no summation over a time grid is involved.

### 2.5 Evaluating aggregate targets

We now express the usual aggregate calibration targets in terms of *p*_*s*_ and *π*_*s*_.

#### 2.5.1 Prevalence rates

Let S(*t*) be the survival function of the model, that is,

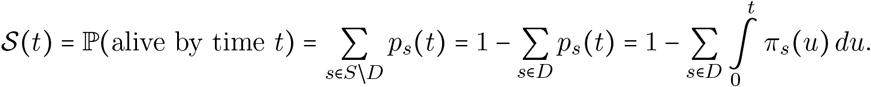

The last expression suits the framework the best, because the transform of 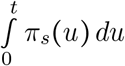 is Π_*s*_ (*ω*)/ *ω*, which is a direct output of the linear system, and accumulating arrivals in death states from time zero greatly reduces the effect of the Gibbs phenomenon.

The prevalence of state *s* at age *t* is the proportion of the survivors who occupy the state:

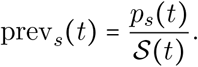

#### 2.5.2 Incidence rates

The instantaneous incidence rate of state *s* is 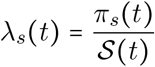. Over an age interval [*t*_1_ , *t*_2_ ], we divide the arrivals in *s* by the person-time lived by survivors,

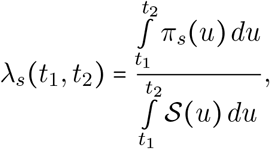

which reduces to the instantaneous rate as *t*_1_ *→ t*_2_. This survivor-weighted rate differs from the naive average 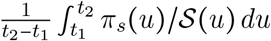, substantially at older ages, where the surviving population falls quickly.

#### 2.5.3 Mortality rates

Mortality rates are the incidence rates of death states, 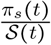 for *s* ∈ *D*, or their interval counterparts. Mortality by cause, and by the state from which death occurs, comes from the death-state equations of (1),

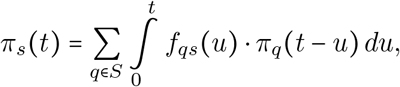

where each term is the flow of deaths *q* → *s*. In frequency space, these terms are 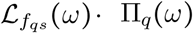, already computed when solving the linear system, so stage-specific mortality targets cost nothing extra.

#### 2.5.4 Incorporating a background mortality rate

Deaths from other causes enter as an independent competing risk with age-dependent hazard *λ*_0_(*t*) and survival 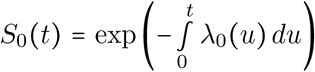, where individuals follow the semi-Markov dynamics until this competing process removes them. Then

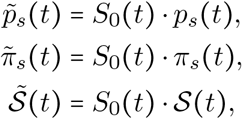

so *S*_0_ (*t*) cancels from the prevalence and from instantaneous incidence rates, but not from interval rates, which become

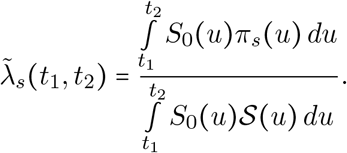

Deaths from other causes while in state *s* occur at density 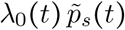, so their rate among survivors is 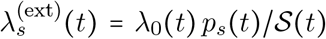, which we evaluate at the start of each one-year age interval.

In our application, we take *S*_0_ from the US life table and set *t* to one in the weights above and in 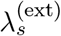; this neglects the cancer’s own share of all-cause mortality and changes no target by more than 0.5% (Supplementary Section 6.1).

#### 2.5.5 Likelihood functions

Consider age intervals 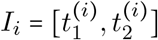 with observed incidence rates 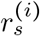 and standard errors 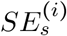, for some state *s*. Assuming each observed rate is normally distributed around the model-predicted rate 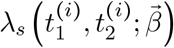 with standard deviation 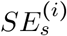, its log-likelihood contribution is

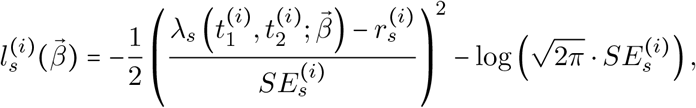

and similarly for prevalence and mortality targets. Assuming independence across targets, the log-likelihood is the sum of these terms; correlated targets would call for a multivariate normal or multinomial likelihood, which requires specifying the correlation structure. Targets with no observed events have no usable standard error under this approximation and are omitted (40 of the 552 targets in our application, all of them preclinical mortality from the cancer or from other causes); a Poisson likelihood would use them.

### 2.6 Including parameter local information

Every quantity in Corollary 2 is an explicit algebraic function of the transformed sub-densities 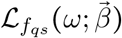, and those are smooth in 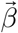 whenever the hazards are, with derivatives obtained by differentiating under the integral sign in the transform. Exact first- and second-order derivatives of the targets are therefore available without finite differencing and, more importantly, without a second pass over the expensive part of the computation.

Differentiating the linear system (2) with respect to a parameter *β*_*k*_ gives

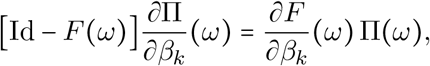

which is the same linear system with a new right-hand side, and similarly at second order, so the factorization computed in step 2 of Algorithm 1 is reused for all *P* gradient components and all *P* (*P* + 1) / 2 Hessian components. The chain rule then propagates these through the target definitions of Section 2.5 and the objective of Section 2.5.5 to give 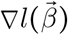 and 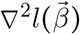 exactly, up to the tolerance of the transform inversion. The Supplementary Material gives the derivations and the resulting expressions in terms of hazards, and Section 3.1 confirms that they are implemented correctly by comparing them with numerical derivatives obtained by perturbing each parameter slightly and re-evaluating the log-likelihood (central finite differences).

### 2.7 Parameter identifiability

The derivatives also support the two standard local identifiability diagnostics. The first is the rank of the sensitivity matrix, the Jacobian of the model-predicted targets with respect to 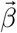. The second is the observed information matrix, the negative Hessian of the log-likelihood (see Section 2.5.5) evaluated at a maximum likelihood estimate, 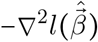. Both are returned by step 6 of Algorithm 1. Eigenvalues at or near zero indicate parameter combinations, given by the corresponding eigenvectors, that the targets do not inform locally [23, 24]. Two cautions apply. First, eigenvalues change with reparameterization, so what counts as “near zero” depends on scaling (we work on the log scale), and a small eigenvalue signals local flatness rather than global non-identifiability.

### 2.8 Parameter estimation leveraging local information

#### 2.8.1 Maximum likelihood estimation

Because exact gradients and Hessians are available, we maximize the log-likelihood with second-order methods from multiple starting points. The likelihood surface of a general semi-Markov model need not be concave and depends on the parameterization, so we use the saddle-free Newton method [25], which reduces to Newton-Raphson where the objective is concave and is not attracted to saddle points elsewhere. Each iteration takes the full saddle-free Newton step, scaled down if it would move any parameter by more than one unit on the log scale, and halved until the objective decreases sufficiently (the Armijo condition). From the estimate of the original analysis, the bladder model converged in eight iterations to a gradient norm of 6.4 × 10^−9^.

#### 2.8.2 Bayesian estimation

Several samplers explore local information, including Hamiltonian Monte Carlo (HMC) [26], the Metropolis-adjusted Langevin algorithm (MALA) [27, 28, 29, 30], and Stein discrepancy methods [31, 32, 33, 34]. We use the Langevin incremental mixture sampler (LIMIS) [35], a modification of the incremental mixture importance sampler of [36], for its balance of effectiveness and simplicity. LIMIS uses the gradient and Hessian of the log-posterior to build a mixture of multivariate Student-*t* approximations (with 3 degrees of freedom), following a deterministic version of the Langevin diffusion. Under a quadratic approximation of the log-posterior, these dynamics have a closed-form solution, whose mean converges to the mode of the approximation and whose covariance converges to the negative inverse Hessian (Supplementary Material, Proposition S1).

### 2.9 Relationship to existing calibration approaches

The framework evaluates a model rather than replacing calibration algorithms. Markov chain Monte Carlo, approximate Bayesian computation, and importance-sampling calibration explore the parameter space, accept aggregate targets, and treat the model as a black box [11, 12, 13]. Any of these methods can call the evaluator described here. The evaluator adds three things. First, accuracy: the only numerical error is inversion tolerance, so no cycle length or random seed needs reporting. Second, cost is independent of target rarity. Several targets are on the order of 10 ^−8^ per person-year, requiring a microsimulation model to simulate enough person-time to resolve them at every evaluation (Section 3.1). Third, differentiability: calibration is usually derivative-free because Monte Carlo objectives lack usable derivatives, not by choice. Exact gradients and Hessians enable Newton-type optimization, gradient-informed sampling, and Section 2.7 diagnostics at only a small multiple of the cost of one evaluation.

The comparison also goes the other way. A deterministic discrete-time cohort model differentiated using automatic differentiation would retain the last two advantages, but it would still have discretization error whose size depends on how the cycle length interacts with the holding-time hazards. Compared with a microsimulation model, the framework forgoes individual histories, heterogeneity, and covariates, features that aggregate targets cannot identify (Section 2.2.1).

### 2.10 Implementation

The open-source package, SMORFS (*Semi-Markov Optimization and Regression by Fourier Series*) implements the framework. The user specifies states and parametric transitions; the package computes the targets and their first and second derivatives, and performs optimization, identifiability analysis, and LIMIS sampling, hiding all frequency-domain calculations. The Supplementary Material lists the specification code for the example model.

## 3 Results

We applied the framework to a continuous-time semi-Markov model of urinary bladder cancer (Figure 2) with a healthy state (*H*), preclinical states (*U*_1_, *U*_2_, *U*_3_) and clinical states (*T*_1_, *T*_2_, *T*_3_) for early, medium, and late stage, and a cancer death state (*D*_*c*_).

**Figure 2:**
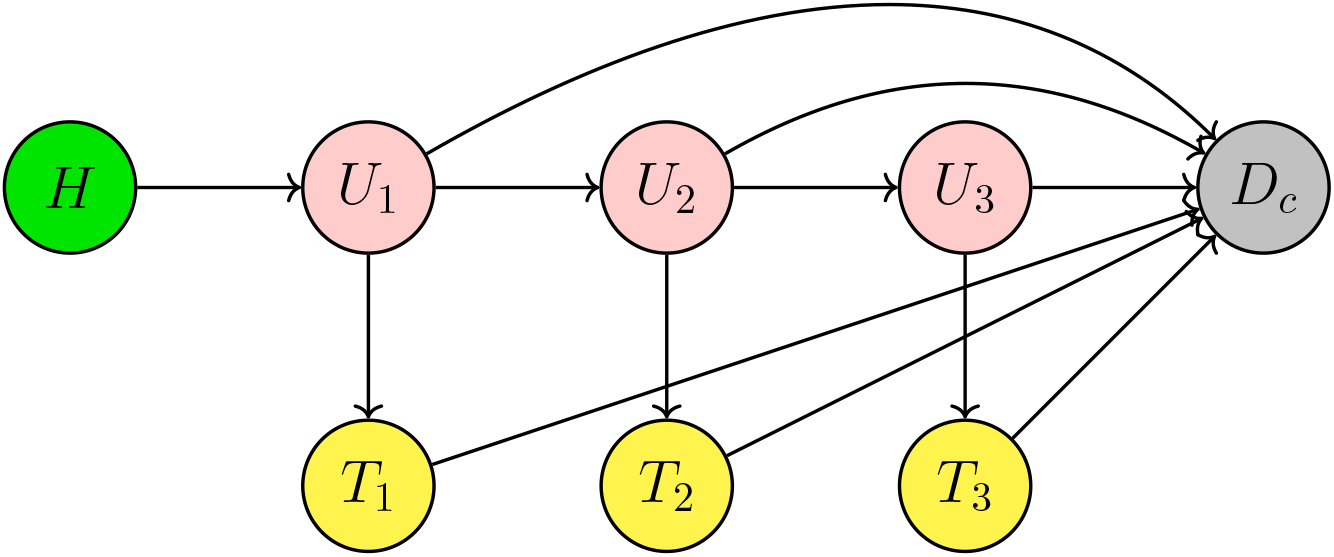
State-transition diagram of the natural history of the urinary bladder cancer semi-Markov model. *H*: healthy, without bladder cancer. *U*_1_, *U*_2_, *U*_3_: preclinical (undiagnosed and untreated) cancer at early, medium, and late stage. *T*_1_, *T*_2_, *T*_3_: clinical (diagnosed and treated) cancer at the same stages. *D*_*c*_: death from bladder cancer. The early, medium, and late stages correspond to the SEER stages in situ or localized, regional, and distant. Arrows show onset (*H* → *U*_1_), progression (*U*_1_ → *U*_2_ → *U*_3_), diagnosis (*U*_*k*_ → *T*_*k*_), and death from the cancer from every preclinical and clinical state. Onset follows a Weibull hazard; every other transition has a constant hazard. Not pictured: death from other causes, a competing risk from every living state.

Onset, *H* → *U*_1_ has a Weibull hazard and all other transitions have constant hazards, resulting in 13 parameters, which we estimate on the log scale for numerical stability. Background mortality was handled as in Section 2.5.4.

We used the SEER data set [37] to construct calibration targets by age and stage: clinical incidence, clinical mortality, preclinical mortality, and preclinical mortality from other causes. Supplementary Section 6 describes the data and reports the rates.

### 3.1 Verification and computational cost

Because every post-onset transition in this model has a constant hazard, its targets can also be computed by convolving the Weibull onset density with a matrix exponential. We implemented this independently, in R and sharing no code with SMORFS, and verified it by step halving to a relative accuracy of 6 × 10^−6^. With background mortality removed, SMORFS reproduces this reference to a maximum relative difference of 7.9 × 10 ^−6^ across the nine target series at ages 35 to 80, that is, to within the reference’s own accuracy, and its pointwise arrival densities agree with a quadrature of the matrix exponential to 1.7 × 10 ^−8^. With background mortality included as in estimation, the analytic gradient agrees with central finite differences of the objective to 8.6 × 10^− 8^ (relative) at a point displaced by 0.1 on the log scale from the maximum likelihood estimate, where the gradient does not vanish, and the analytic Hessian agrees with central differences of an accurate gradient to 9.7 × 10 ^−7^ at the estimate, where it is positive definite.

Table 3 compares four ways of evaluating the same targets. A discrete-time cohort approximation of the same model deviates from the reference by up to 3.3% with a one-month cycle and 42% with a one-year cycle, because the mean holding times in *U*_1_ and *U*_2_ at the parameter values used are 0.34 and 0.27 years, shorter than the cycles. A microsimulation of two million individuals deviates by up to 811%, with the largest errors in the sparsest targets. SMORFS evaluates all targets in 1.3 milliseconds, their gradient in 2.5 milliseconds, and their Hessian in 9.1 milliseconds: about 11 times faster than the reference (14 milliseconds) and five times faster than the one-month cohort approximation (6 milliseconds), which is four thousand times less accurate. Only the one-year cohort approximation is faster (1 millisecond), at an error of 42%. Only the SMORFS library has been optimized, so these timings show that the method is practical, not that it is faster in general. The reference exists only because the post-onset holding times are exponential; when they are not, no such shortcut is available, while the frequency-domain evaluation applies unchanged.

### 3.2 Maximum likelihood estimation

We started Newton’s method, with the analytic gradient and Hessian, from 21 points: the estimate of the original analysis and 20 points drawn uniformly within one unit of the maximum likelihood estimate on the log scale. With full steps, 14 runs, including the one from the original estimate, converged to the same optimum; six of the other seven did so with steps capped at one log unit, and one ended at a degenerate point. No other interior optimum was found. Figure 3 shows the fit at the optimum; the observed information there gives Table 2’s confidence intervals.

**Table 2:**
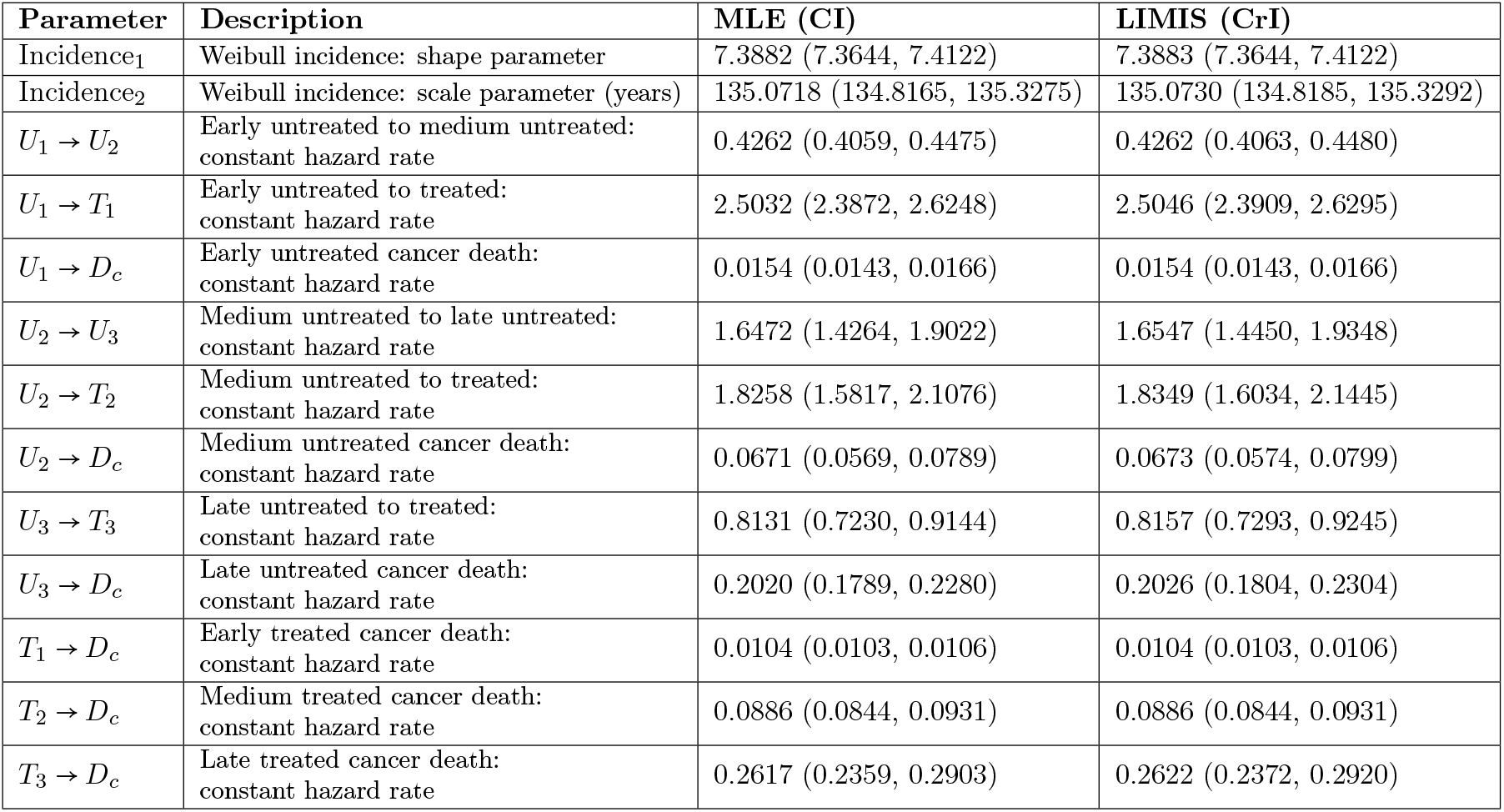
Model parameters: maximum likelihood estimates with 95% confidence intervals (CI) from the observed information, 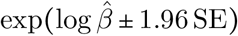, and posterior medians with 95% credible intervals (CrI), the 2.5% and 97.5% weighted quantiles of the 20 pooled LIMIS runs. Rates are per year; values are rounded to four decimal places. MLE: maximum likelihood estimation; LIMIS: Langevin incremental mixture importance sampling.

| Parameter | Description | MLE (CI) | LIMIS (CrI) |
| --- | --- | --- | --- |
| Incidence<sub>1</sub> | Weibull incidence: shape parameter | 7.3882 (7.3644, 7.4122) | 7.3883 (7.3644, 7.4122) |
| Incidence<sub>2</sub> | Weibull incidence: scale parameter (years) | 135.0718 (134.8165, 135.3275) | 135.0730 (134.8185, 135.3292) |
| $U_1 \rightarrow U_2$ | Early untreated to medium untreated:<br>constant hazard rate | 0.4262 (0.4059, 0.4475) | 0.4262 (0.4063, 0.4480) |
| $U_1 \rightarrow T_1$ | Early untreated to treated:<br>constant hazard rate | 2.5032 (2.3872, 2.6248) | 2.5046 (2.3909, 2.6295) |
| $U_1 \rightarrow D_c$ | Early untreated cancer death:<br>constant hazard rate | 0.0154 (0.0143, 0.0166) | 0.0154 (0.0143, 0.0166) |
| $U_2 \rightarrow U_3$ | Medium untreated to late untreated:<br>constant hazard rate | 1.6472 (1.4264, 1.9022) | 1.6547 (1.4450, 1.9348) |
| $U_2 \rightarrow T_2$ | Medium untreated to treated:<br>constant hazard rate | 1.8258 (1.5817, 2.1076) | 1.8349 (1.6034, 2.1445) |
| $U_2 \rightarrow D_c$ | Medium untreated cancer death:<br>constant hazard rate | 0.0671 (0.0569, 0.0789) | 0.0673 (0.0574, 0.0799) |
| $U_3 \rightarrow T_3$ | Late untreated to treated:<br>constant hazard rate | 0.8131 (0.7230, 0.9144) | 0.8157 (0.7293, 0.9245) |
| $U_3 \rightarrow D_c$ | Late untreated cancer death:<br>constant hazard rate | 0.2020 (0.1789, 0.2280) | 0.2026 (0.1804, 0.2304) |
| $T_1 \rightarrow D_c$ | Early treated cancer death:<br>constant hazard rate | 0.0104 (0.0103, 0.0106) | 0.0104 (0.0103, 0.0106) |
| $T_2 \rightarrow D_c$ | Medium treated cancer death:<br>constant hazard rate | 0.0886 (0.0844, 0.0931) | 0.0886 (0.0844, 0.0931) |
| $T_3 \rightarrow D_c$ | Late treated cancer death:<br>constant hazard rate | 0.2617 (0.2359, 0.2903) | 0.2622 (0.2372, 0.2920) |

**Table 3:**
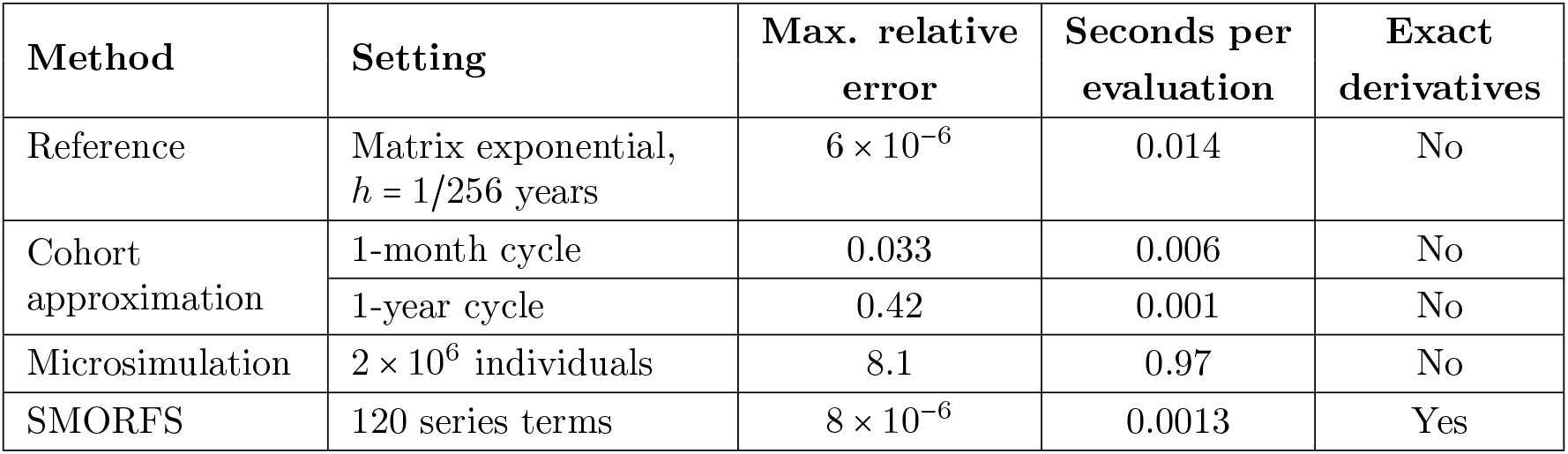
Accuracy and cost of four ways of evaluating the same calibration targets for the bladder cancer model, with background mortality removed, at the parameter values of Table 2 of the original analysis. Errors are relative to the reference implementation, maximized over the nine target series at ages 35 to 80; for the reference itself, the error is the change from halving the time step, so SMORFS agrees with it to within its accuracy. SMORFS’s gradient and Hessian take 0.0025 and 0.0091 seconds. All methods are single-threaded; times are medians of 10 runs on an Apple M4 Max (16 cores, 128 GB; macOS 26.6.2, Python 3.12.15, R 4.5.3).

| Method | Setting | Max. relative error | Seconds per evaluation | Exact derivatives |
| --- | --- | --- | --- | --- |
| Reference | Matrix exponential,<br>$h = 1/256$ years | $6 \times 10^{-6}$ | 0.014 | No |
| Cohort approximation | 1-month cycle | 0.033 | 0.006 | No |
|  | 1-year cycle | 0.42 | 0.001 | No |
| Microsimulation | $2 \times 10^6$ individuals | 8.1 | 0.97 | No |
| SMORFS | 120 series terms | $8 \times 10^{-6}$ | 0.0013 | Yes |

**Figure 3:**
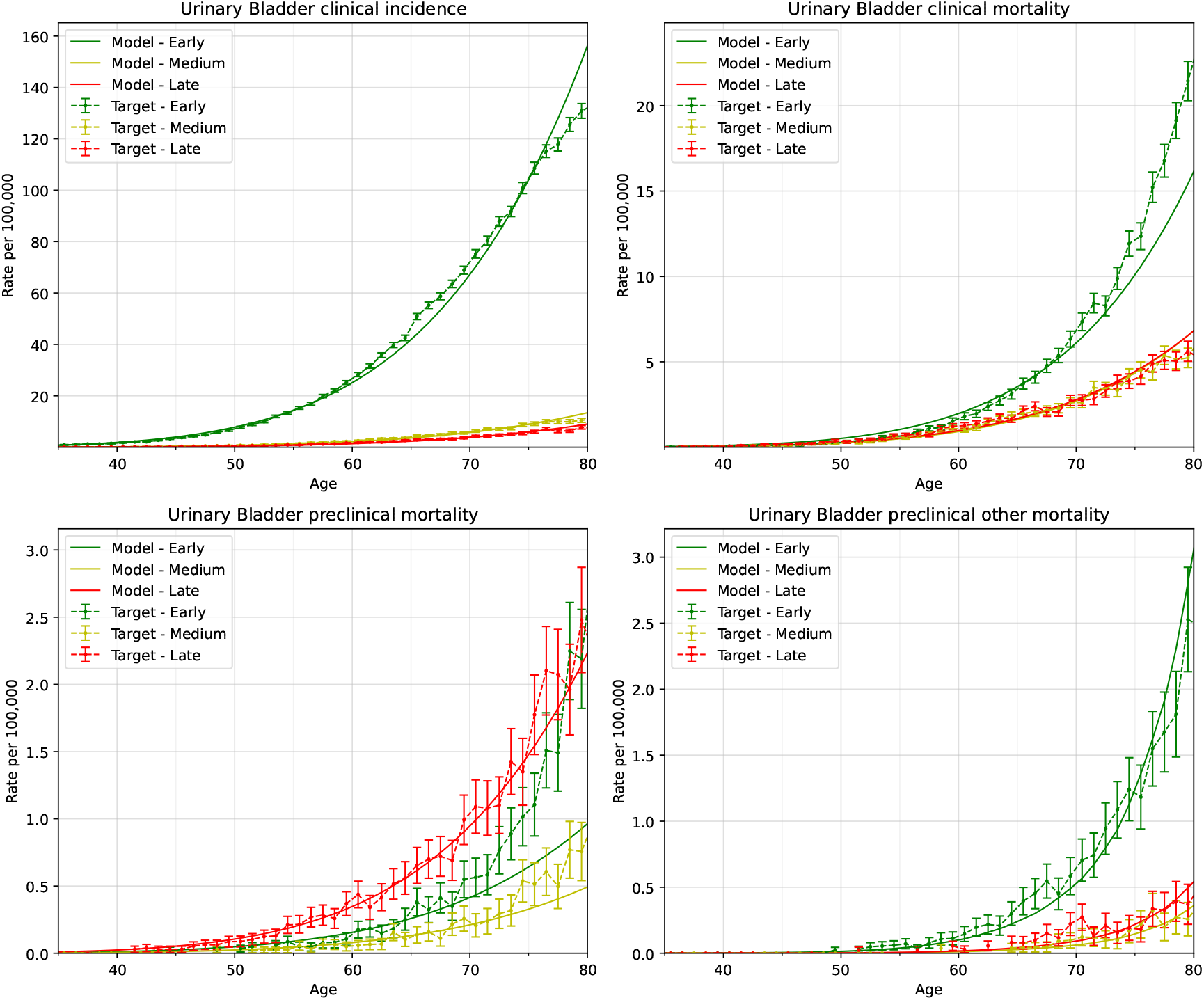
Comparison between model-predicted outputs evaluated at the maximum like-lihood estimate (MLE) and calibration targets. Error bars show calibration targets with 95% confidence intervals (CI). The solid line shows the outputs generated by the model evaluated at the MLE. Targets based on 1 to 4 events are not shown, as the SEER data-use agreement requires; they are used in the calibration.

The eigenvalues of the observed information range from 60 to 1.9 × 10^7^, a condition number of 3 × 10^5^. The eigenvectors of the two smallest load almost equally on all exits from *U*_2_ and on both exits from *U*_3_: the targets determine where individuals go from each preclinical state far better than how long they stay. At the estimate, the mean preclinical holding times are 0.34, 0.28, and 0.99 years in *U*_1_, *U*_2_, and *U*_3_.

The fit is systematically off for the early stage. With standardized residuals *z* = model − target /*SE*, the 512 targets with at least one event give ∑*z*^2^ = 4,363, or 8.5 per target, against about 1 for a model fitting within sampling error. Early-stage clinical incidence and mortality contribute 63% of this total from 17% of the targets. Modeled early-stage incidence lies below the targets from about age 57 to 75 (*z* = −11 at 65) and above them after age 75 (*z* = +19 at 80), and early-stage clinical mortality shows the opposite pattern, from *z* = + 3 at 60 to *z* = − 11 at 80. The misfit belongs to the model, not to its evaluation: a cohort or microsimulation version of the same model computes the same targets (Table 3) and would fit no better. The confidence and credible intervals of Table 2 therefore reflect sampling error in the targets, conditional on the model structure, and not the structural error; a simple overdispersion correction would widen them by a factor of about 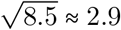.

### 3.3 Bayesian estimation

For Bayesian estimation we used uniform priors on the log-scale parameters, centered at the maximum likelihood estimate, with half-width equal to the larger of 0.3 (a factor of 1.35) and four standard errors from the observed information. These bounds are computational support for the sampler, chosen to contain the posterior, rather than substantive prior beliefs; because they are centered on the estimate, the credible intervals should be read as conditional on them. Figure 4 shows that the 95% prior predictive band covers every clinical target and every target with two or more events; the only targets outside it are the 40 preclinical targets with no events, which carry no weight in the likelihood (Section 2.5.5), and two single-event targets just above it.

**Figure 4:**
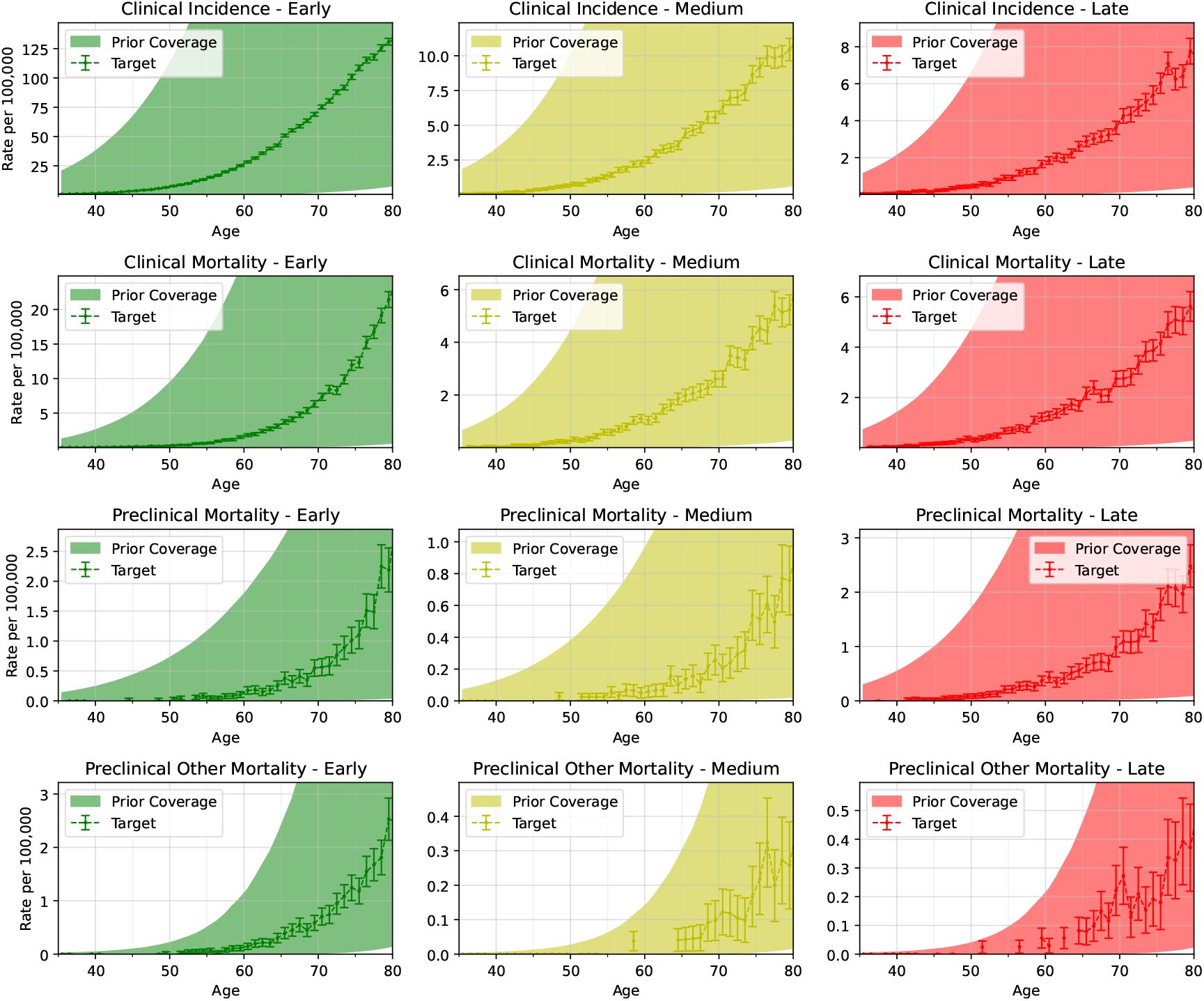
Prior predictive coverage: 95% intervals of model-predicted targets under the prior (shaded), with the calibration targets and their 95% confidence intervals. Targets based on 1 to 4 events are not shown, as the SEER data-use agreement requires; they are used in the calibration.

We ran 20 independent executions of LIMIS, each with 10,000 initial samples and 100 mixture steps of 1,000 samples, for 2,200,000 samples in total; posterior summaries are weighted quantiles of the pooled samples. The pooled effective sample size was about 1.08 million, the 20 runs’ posterior means agreed to within 0.006 posterior standard deviations, the posterior standard deviations were 1.00 to 1.01 times the information-matrix standard errors, and no marginal placed more than 0.06% of its mass near a bound. With bounds of four standard errors instead, the posterior medians and 95% credible limits changed by at most 0.005 posterior standard deviations. Exits from the same preclinical state were positively correlated in the posterior, from 0.62 (*U*_1_ → *U*_2_ with *U*_1_ → *D*_*c*_) to 0.99 (*U*_2_ → *U*_3_ with *U*_2_ → *T*_2_): the uncertainty lies mostly along directions that scale all exits from a state together, changing how long individuals stay but not where they go, as in Section 3.2. The Weibull shape and scale were correlated at 0.95. Figures 5 to 7 show the fit, the marginal densities, and the correlogram.

**Figure 5:**
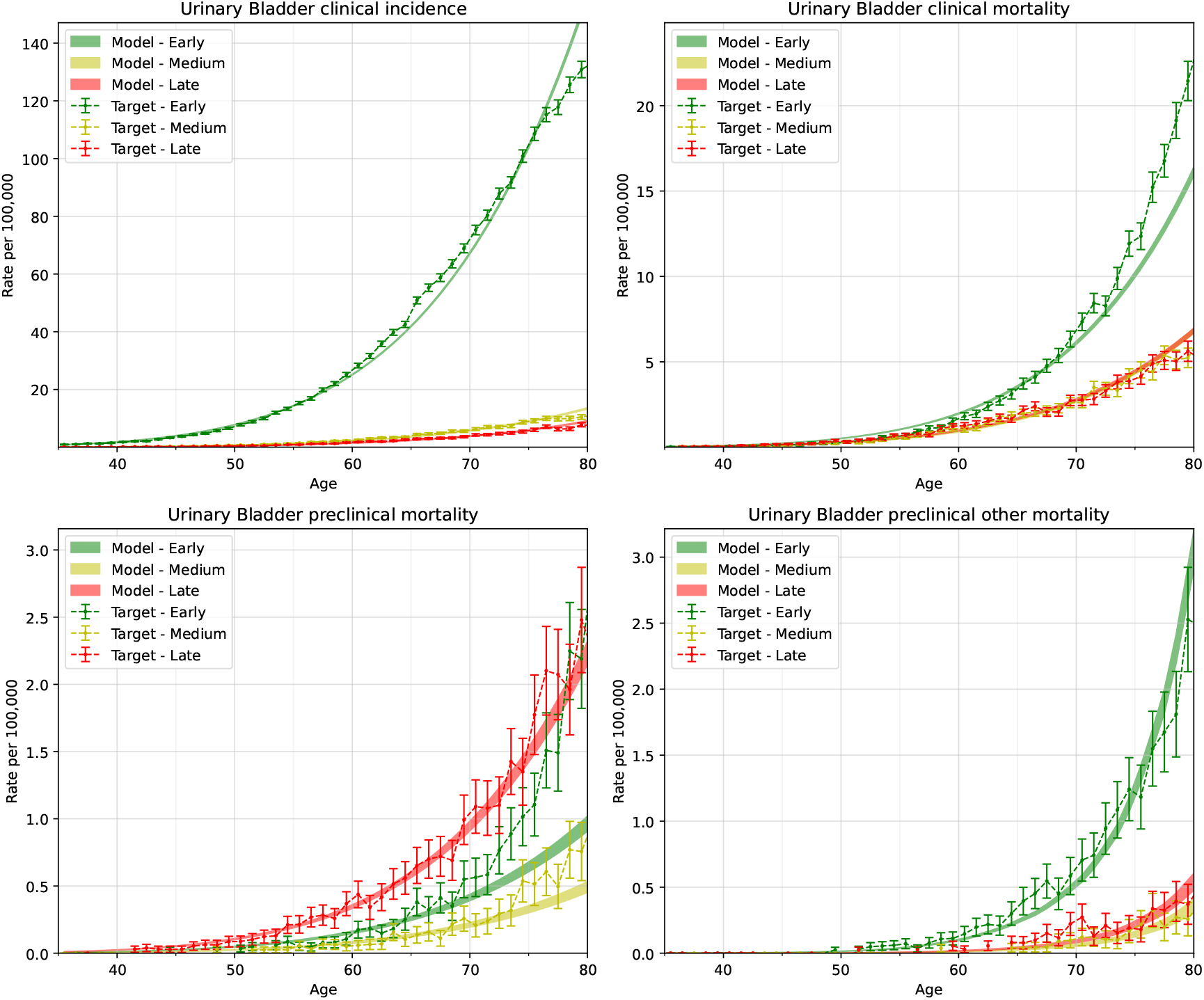
Comparison between posterior model-predicted outputs and calibration targets. Error bars show calibration targets with their 95% confidence intervals (CI). The shaded area represents the 95% posterior model-predictive interval of the outcomes computed from 10,000 parameter draws resampled from the pooled LIMIS samples in proportion to their importance weights. Targets based on 1 to 4 events are not shown, as the SEER data-use agreement requires; they are used in the calibration.

**Figure 6:**
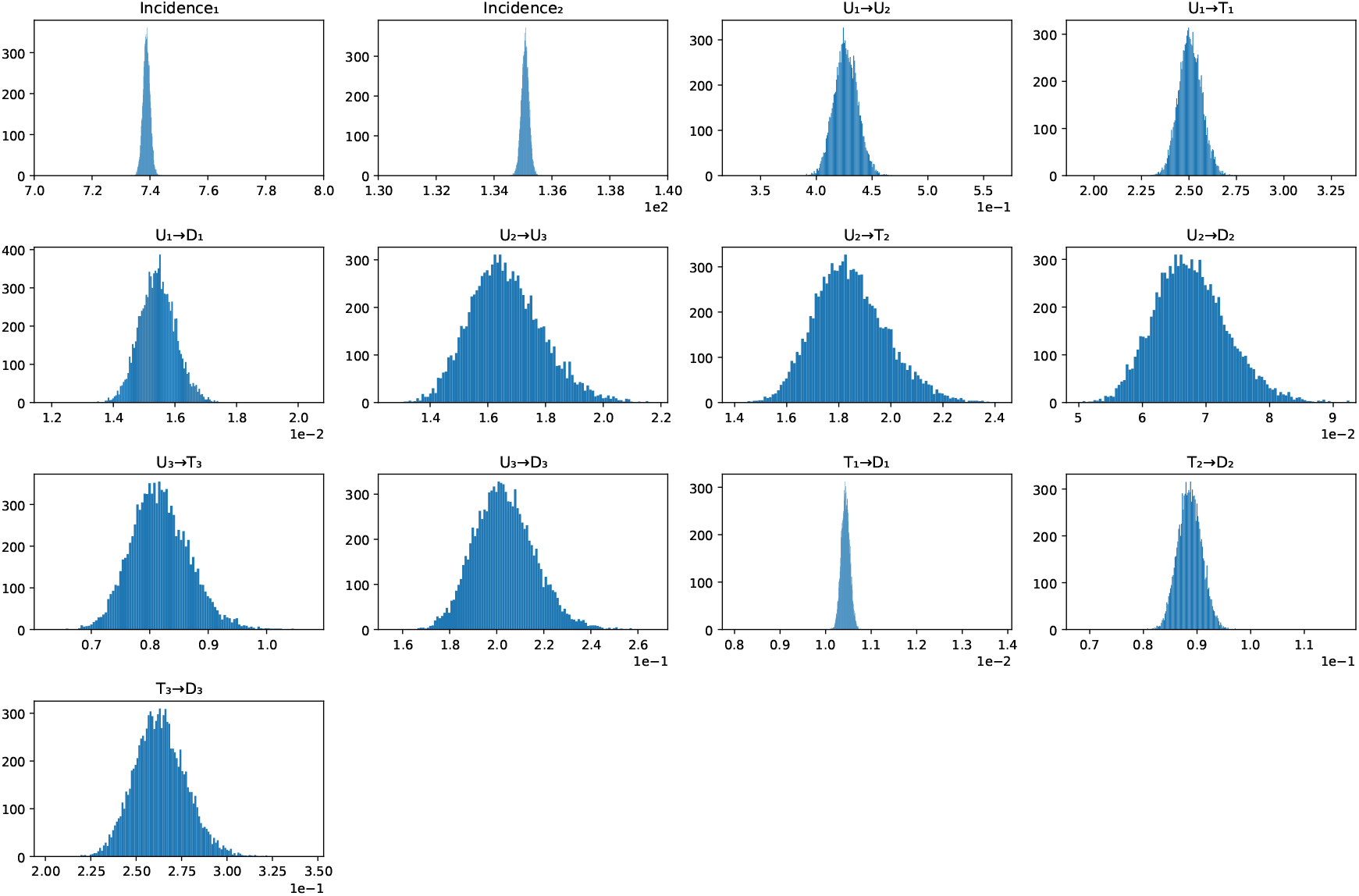
Posterior marginal densities of estimated parameters drawn from the Langevin incremental mixture importance sampling (LIMIS) algorithm. The x-axis of each panel spans the bounds of its uniform prior, so the width of each histogram relative to its axis shows how much the data narrowed the prior; the exceptions are the Weibull shape and scale (red axes), shown on 7 to 8 and 130 to 140 years so that their posteriors are visible, whose prior bounds are 5.47 to 9.97 and 100.1 to 182.3 years.

**Figure 7:**
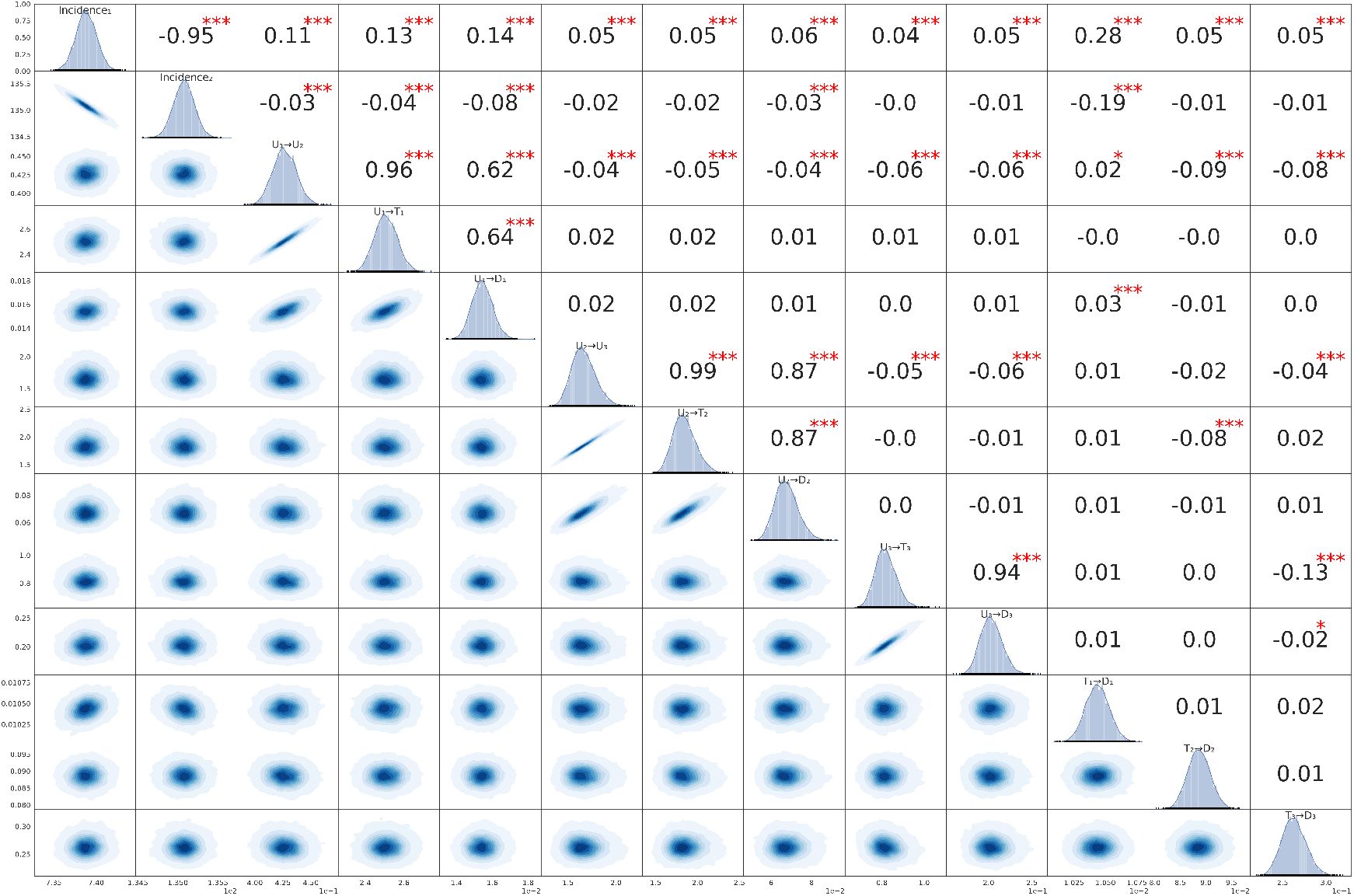
Scatter plot of pairs of estimated model parameters with correlation coefficient and posterior marginal distributions.

Table 2 shows the resulting 95% credible intervals for the model parameters in the LIMIS column, while Figure 5 shows the 95% posterior model-predictive intervals for the targets.

## 4 Discussion

We developed a framework for evaluating and estimating continuous-time semi-Markov natural history models from aggregate data by working in the frequency domain. We used it to calibrate a urinary bladder cancer model against SEER targets, applying both MLE and Bayesian inference with the LIMIS algorithm. It reproduced an independent implementation to within that implementation’s own accuracy and showed which parameter combinations the aggregate targets leave weakly identified, but the model did not reproduce the age gradient of early-stage incidence and mortality.

The estimation literature for multi-state survival and semi-Markov models often requires individual-level time-to-event data [2, 3, 9], while calibration methods for aggregate targets treat the model as a black box [11, 12, 13]. Our contribution bridges the two by providing a deterministic, differentiable evaluator that supplies the targets those methods compare against, as well as their derivatives.

The principal limitation is structural. Hazards may depend on the time spent in a state rather than on attained age or calendar time. In our example, the age gradient in incidence comes entirely from the Weibull onset hazard, where local and global time coincide; progression after onset therefore cannot vary with age. These constraints are a plausible source of the misfit of Section 3.2, as is the restriction that the death rate after an early-stage diagnosis cannot depend on the age at diagnosis. Because all post-onset holding times are exponential, the example is also close to a Markov model. We chose it because it allows the independent check described in Section 3.1; the framework’s advantage is greater when holding times are not exponential. Models that require both time scales, such as those with age-dependent progression or screening at calendar times, require the more general and costly formulation of [38]. Aggregate data also introduce ecological bias and unobserved heterogeneity, and without screening data they leave preclinical holding times weakly identified. Clinical mortality targets omit deaths of cases diagnosed before 2004 and are biased downward (Supplementary Section 6). Parametric hazards may be misspecified, the normal likelihood is a poor approximation for the 93 preclinical targets with three or fewer events, background mortality is assumed to be independent of the disease process, and no target was held out. Therefore, Figures 3 and 5 assess calibration rather than external validity.

Natural extensions include spline-based hazards, mixtures for latent heterogeneity, and screening and treatment modules built on the calibrated natural history.

Working in the frequency domain makes continuous-time semi-Markov natural history models cheap enough to evaluate repeatedly and smooth enough to differentiate, which turns their calibration to aggregate data from a search into an estimation problem. The software is openly available at https://github.com/fsstol/SMORFS (release v1.0.1).

## Acknowledgements

This study used data from the Surveillance, Epidemiology, and End Results (SEER) Program of the National Cancer Institute: Incidence – SEER Research Data, 17 Registries, Nov 2023 Sub (2000–2021), and Incidence-Based Mortality – SEER Research Data, 17 Registries, Nov 2023 Sub (2000–2021). The interpretation and reporting of these data are the sole responsibility of the authors.

The authors used generative AI tools (Claude, Anthropic, including Claude Code) to review and debug the software, write tests and verification scripts, and draft and edit the revised manuscript and the response to reviewers. The authors reviewed and verified all code, analyses, and text, and take full responsibility for them.

## Statements and Declarations

### Ethical considerations

This study analyzed de-identified, aggregate data from the SEER Program, obtained under the SEER Research Data Use Agreement. It involved no human participants and no identifiable private information, so institutional review board approval was not required.

### Consent to participate

Not applicable.

### Consent for publication

Not applicable.

### Data availability

The calibration targets were constructed from SEER data, which are available from the National Cancer Institute under the SEER Research Data Use Agreement; the agreement does not permit the authors to redistribute them. The software, the parameter estimates, instructions for rebuilding the targets with SEER*Stat, and a synthetic data set simulated from the fitted model, on which every step of the analysis can be run, are available at https://github.com/fsstol/SMORFS (release v1.0.1). The life table used for background mortality is publicly available from the US Social Security Administration.

