## Supplementary Material for "Frequency-Domain Evaluation and Estimation of Semi-Markov Disease Natural History Models"

#### 1 Proof of Proposition 1

**Proposition 1.** The densities $(\pi_s)_{s\in S}$ of Definition 1 in the main text are the unique solution of the system of convolution equations

$$\pi_s(t) = \mathbb{1}_{\{s=s_0\}} \cdot \delta_0(t) + \sum_{q\in S} \int_0^t f_{qs}(u) \cdot \pi_q(t-u)\, du \quad \forall s,\ t \geq 0, \tag{1}$$

where $\delta_0$ is the Dirac delta distribution at time 0. For each state $s \in S$, the function $g_s$ is equal to the convolution of the distribution $\pi_s(t)$ and the function $d_s(t) := \mathbb{E}(\min(\phi_s, t))$.

*Proof.* We form a matrix of functions $(g_{sr}(t))_{s,r\in S}$ such that for each pair of states $s, r \in S$, $g_{sr}(t)$ is defined as the expectation of the random variable $I_{sr}(t)$ quantifying the time spent in state $r$ when starting from state $s$ and with a remaining observation time of $t$ (and $I_{sr}(t) \equiv 0$ for $t \leq 0$).

From this definition of $I_{sr}(t)$ we have:

$$I_{sr}(t) = \mathbb{1}_{\{s=r\}} \cdot \min(\phi_s, t) + \sum_{q\in S} \mathbb{1}_{\{s\to q\}} I_{qr}(t-\phi_s),$$

since the first term contributes to the observed time from staying at $s$ for the random time $\phi_s$ when the remaining observation time is $t$ and the initial and observation states $s, r$ coincide. The second sum comes from partitioning the process over the next visited state $q \in S$ and considering the contribution of the remaining observation time $t - \phi_s$ starting from the new state $q$. Then, by taking expectations, we obtain

$$g_{sr}(t) = \mathbb{E}(I_{sr}(t)) = \mathbb{1}_{\{s=r\}} \cdot \mathbb{E}(\min(\phi_s, t)) + \sum_{q\in S} \mathbb{E}(\mathbb{1}_{\{s\to q\}} I_{qr}(t-\phi_s)).$$

By using the density $f_s$ of $\phi_s$ to condition on its value, each term of the sum can be expressed as

$$\mathbb{E}(\mathbb{1}_{\{s\to q\}} \cdot I_{qr}(t-\phi_s)) = \int_0^t \mathbb{E}(\mathbb{1}_{\{s\to q\}} \cdot I_{qr}(t-u) | \phi_s = u) f_s(u)\, du,$$

which by definition of the marginal density functions $f_{sq}$ and independence of the remainder of the process after transitioning to $q$ is also equal to

$$\mathbb{E}(\mathbb{1}_{\{s\to q\}} \cdot I_{qr}(t-\phi_s)) = \int_0^t \mathbb{E}(I_{qr}(t-u)) f_{sq}(u)\, du = \int_0^t f_{sq}(u) \cdot g_{qr}(t-u)\, du.$$

We have thus shown that the matrix of functions $g$ satisfies the matrix convolution system

$$g = d + f * g \qquad \Longleftrightarrow \qquad g = (\mathbf{Id} - f)^{-1} * d,$$

over states $S$, where $\mathbf{Id} = \mathbf{Id}_S$ is the identity distribution (holding the standard Dirac delta distribution in its diagonal components and zero elsewhere), $d$ is a diagonal matrix of functions $d_s(t) = \mathbb{E}(\min(\phi_s, t))$ and $f$ is the matrix of functions $f_{sq}(t)$. Since we are interested in the functions $g_{s_0 s}$ obtained when starting from $s_0$, we can take the canonical column vector of distributions $e_{s_0}$ such that it holds the Dirac delta distribution as its $s_0$-th component and zero for the rest, and write

$$g_{s_0,\cdot} = e_{s_0}^\top * (\mathbf{Id} - f)^{-1} * d,$$

If we define the column vector of distributions $\pi$ as the transpose of the first convolution, then it is such that $(\mathbf{Id} - h^\top) * \pi = e_{s_0}$, or $\pi = e_{s_0} + h^\top * \pi$, which is readily seen to be equivalent to

$$\pi_s(t) = \mathbb{1}_{\{s=s_0\}} \cdot \delta_0(t) + \sum_{q \in S} \int_0^t f_{qs}(u) \cdot \pi_q(t-u)\, du \quad \forall s \in S,\ t \geq 0,$$

which is the claimed system for the distributions $(\pi_s)_{s \in S}$. Finally, noting that

$$g_s(t) = g_{s_0 s}(t) = (\pi^\top * d)_s(t) = (\pi_s * d_s)(t),$$

we recover the functions $(g_s(t))_{s \in S}$ as claimed.

It remains to identify this $\pi$ with the arrival densities of Definition 1, since the construction above defines $\pi$ through $g$. Let $m_s(t) \coloneqq \mathbb{E}(N_s(t))$. An arrival in $s$ during $[0,t]$ is either the initial placement (if $s = s_0$) or a transition $q \to s$ that ends a visit to some state $q$ begun at an earlier arrival. Because the holding time and destination after each arrival are independent of the past, with sub-densities $f_{qs}$,

$$m_s(t) = \mathbb{1}_{\{s=s_0\}} + \sum_{q \in S} \int_0^t \Big( \int_0^{t-u} f_{qs}(v)\, dv \Big)\, dm_q(u),$$

the Markov renewal equation of [1]. Differentiating in $t$ in the sense of distributions shows that $\pi_s = dm_s/dt$ satisfies (1). Since (1) has a unique solution when $S$ is finite ([1], Corollary 3.16), the arrival densities coincide with the $\pi$ constructed above, which proves both claims. □

#### 2 Proof of Corollary 2

**Corollary 2.** The functions $(\Pi_s(\omega))_{s \in S}$ satisfy the system of equations

$$\Pi_s(\omega) = \mathbb{1}_{\{s=s_0\}} + \sum_{q \in S} \mathcal{L}_{f_{qs}}(\omega) \cdot \Pi_q(\omega)$$

for any $\omega \in \mathbb{C}$ such that the Laplace transforms exist and the system has a unique solution. For any $s \in S$,

$$\mathcal{L}_{g_s}(\omega) = \mathcal{L}_{d_s}(\omega) \cdot \Pi_s(\omega),$$

for any $\omega \in \mathbb{C}$ such that the involved quantities exist uniquely.

The Laplace transform of $d_s$ has a closed-form expression in terms of that of $f_s$:

$$\mathcal{L}_{d_s}(\omega) = \frac{1}{\omega^2} \cdot \left(1 - \mathcal{L}_{f_s}(\omega)\right).$$

*Proof.* The first and second claims follow from taking the Laplace transforms in Proposition 1, identifying $\Pi_s = \mathcal{L}_{\pi_s}$ and using the convolution property.

For the third claim, note that

$$d_s(t) = \mathbb{E}(\min(\phi_s, t)) = \int_0^t u \cdot f_s(u)\, du + t \cdot \int_t^\infty f_s(u)\, du$$

$$\implies d_s'(t) = t \cdot f_s(t) + \int_t^\infty f_s(u)\, du - t \cdot f_s(t) = \int_t^\infty f_s(u)\, du = 1 - F_{\phi_s}(t).$$

Using the fact that $d_s(0) = 0$ and the derivative and primitive properties of the Laplace transform, we obtain

$$\omega \cdot \mathcal{L}_{d_s}(\omega) = \mathcal{L}_{d_s'}(\omega) = \mathcal{L}_1(\omega) - \frac{1}{\omega}\mathcal{L}_{f_s}(\omega) = \frac{1 - \mathcal{L}_{f_s}(\omega)}{\omega},$$

which implies the claim. □

### 3 Proof of Corollary 3

**Corollary 3.** The population profile functions are obtained as convolutions of each state's arrival distribution and the survival functions of their holding times:

$$p_s(t) = \mathbb{P}(X_t = s) = (\pi_s \star S_s)(t) \quad \forall s \in S.$$

The expected discounted occupation times over an infinite time horizon have explicit expressions in terms of the Laplace transforms of the holding times and the arrival time distribution functions in frequency space:

$$\mathbb{E}\left(\int_0^\infty \mathbb{1}_{\{X_t = s\}} \cdot e^{-\alpha t}\, dt\right) = \frac{1 - \mathcal{L}_{f_s}(\alpha)}{\alpha} \cdot \Pi_s(\alpha),$$

for any $\alpha \in \mathbb{R}$ such that these transforms exist.

*Proof.* For the first claim, fix some state $s \in S$ and note that for any $t > 0$,

$$p_s(t) = \mathbb{P}(X_t = s) = \frac{d}{dt}\mathbb{E}\left(\int_0^t \mathbb{1}_{\{X_t = s\}}\, dt\right) = g_s'(t)$$

$$\implies \mathcal{L}_{p_s}(\omega) = \omega \cdot \mathcal{L}_{g_s}(\omega) = \omega \cdot \mathcal{L}_{d_s}(\omega) \cdot \Pi_s(\omega) = \frac{1 - \mathcal{L}_{f_s}(\omega)}{\omega} \cdot \Pi_s(\omega),$$

where the equalities in the second line follow from the derivative property of Laplace transforms and Corollary 2. As we already saw in that proof, $\frac{1 - \mathcal{L}_{f_s}(\omega)}{\omega} = \mathcal{L}_{S_s}(\omega)$ the Laplace transform of state $s$'s holding time survival function, and so we deduce that

$$\mathcal{L}_{p_s}(\omega) = \mathcal{L}_{S_s}(\omega) \cdot \Pi_s(\omega),$$

from which we conclude from the convolution and uniqueness properties.

For the second claim, evaluate this transform at the real frequency $\omega = \alpha > 0$ to obtain

$$\mathbb{E}\left(\int_0^\infty \mathbb{1}_{\{X_t = s\}} \cdot e^{-\alpha t}\, dt\right) = \mathcal{L}_{p_s}(\alpha) = \frac{1 - \mathcal{L}_{f_s}(\alpha)}{\alpha} \cdot \Pi_s(\alpha),$$

as claimed. □

*Remark.* The restriction to $\alpha > 0$ is what makes the infinite-horizon expression finite. It is not a limitation in practice: the undiscounted lifetime occupation time of a transient state is the limit $\alpha \to 0^+$, or directly $g_s(\infty)$.

### 4 Langevin dynamics under a quadratic approximation

The LIMIS sampler of the main text (Section 2.8.2) follows a deterministic version of the Langevin diffusion. The following result, cited in the main text, gives its solution when the log-posterior is replaced by a quadratic approximation.

*Proposition* S1. Let $l(\vec{\beta})$ denote the log-posterior (and with a flat prior, the log-likelihood) evaluated at $\vec{\beta}$, and consider the continuous-time, deterministic Langevin system

$$\begin{aligned}
\vec{\beta}'(t) &= \frac{1}{2}\nabla l(\beta(t)), \\
\Sigma'(t) &= \frac{1}{2}\nabla^2 l(\beta(t)) \cdot \Sigma(t) + \Sigma(t) \cdot \frac{1}{2}\nabla^2 l(\beta(t)) + \mathrm{Id}, \\
& \vec{\beta}(0) = \vec{\beta}_0, \quad \Sigma(0) = \Sigma_0.
\end{aligned}$$

If $l$ is replaced by its second-order approximation around the initial condition,

$$l(\vec{\beta}) = l(\vec{\beta}_0) + \nabla l(\vec{\beta}_0)^\top \cdot (\vec{\beta} - \vec{\beta}_0) + \frac{1}{2}(\vec{\beta} - \vec{\beta}_0)^\top \cdot \nabla^2 l(\vec{\beta}_0) \cdot (\vec{\beta} - \vec{\beta}_0),$$

then we can obtain analytical solutions to the Langevin system, namely

$$\begin{aligned}
\vec{\beta}(t) &= \vec{\beta}_0 + (e^{\frac{1}{2}Ht} - \mathrm{Id})H^{-1}\vec{J}, \\
\Sigma(t) &= -H^{-1} + e^{\frac{1}{2}Ht} \cdot \left(\Sigma_0 + H^{-1}\right) \cdot e^{\frac{1}{2}Ht},
\end{aligned}$$

where $\vec{J} := \nabla l(\vec{\beta}_0)$, $H := \nabla^2 l(\vec{\beta}_0)$, provided that the Hessian matrix $H$ is invertible. If $H$ is negative definite, then as $t \to \infty$, $\vec{\beta}(t) \to \vec{\beta}_0 - H^{-1}\vec{J}$, the mode of the quadratic approximation, and $\Sigma(t) \to -H^{-1}$.

*Proof.* For the second-order approximation of $l$ and the given definitions of $\vec{J}$ and $H$, the Langevin system reads

$$\begin{aligned}
\vec{\beta}'(t) &= \frac{1}{2}\nabla l(\vec{\beta}(t)) = \frac{1}{2}\nabla l(\vec{\beta}_0) + \frac{1}{2}\nabla^2 l(\vec{\beta}_0) \cdot (\vec{\beta}(t) - \vec{\beta}_0) = \frac{1}{2}\vec{J} + \frac{1}{2}H \cdot (\vec{\beta}(t) - \vec{\beta}_0), \\
\Sigma'(t) &= \frac{1}{2}\nabla^2 l(\vec{\beta}(t)) \cdot \Sigma(t) + \Sigma(t) \cdot \frac{1}{2}\nabla^2 l(\vec{\beta}(t)) + \mathrm{Id} = \frac{1}{2}H \cdot \Sigma(t) + \Sigma(t) \cdot \frac{1}{2}H + \mathrm{Id}, \\
& \vec{\beta}(0) = \vec{\beta}_0, \quad \Sigma(0) = \Sigma_0.
\end{aligned}$$

By making the affine change of variables $\tilde{\beta}(t) := \vec{\beta}(t) - \vec{\beta}_0$, $\tilde{\Sigma}(t) := \Sigma(t) + H^{-1}$, this becomes

$$\begin{aligned}
\tilde{\beta}'(t) &= \frac{1}{2}\vec{J} + \frac{1}{2}H \cdot \tilde{\beta}(t), \\
\tilde{\Sigma}'(t) &= \frac{1}{2}H \cdot \tilde{\Sigma}(t) + \tilde{\Sigma}(t) \cdot \frac{1}{2}H, \\
& \tilde{\beta}(0) = \vec{0}, \quad \tilde{\Sigma}(0) = \Sigma_0 + H^{-1} =: \tilde{\Sigma}_0,
\end{aligned}$$

from which it is easy to check that the solutions are $\tilde{\beta}(t) = (e^{\frac{1}{2}Ht} - \mathrm{Id})H^{-1}\vec{J}$, $\tilde{\Sigma}(t) = e^{\frac{1}{2}Ht} \cdot \tilde{\Sigma}_0 \cdot e^{\frac{1}{2}Ht}$, using the fact that $\frac{d}{dt}e^{\frac{1}{2}Ht} = \frac{1}{2}H \cdot e^{\frac{1}{2}Ht} = e^{\frac{1}{2}Ht} \cdot \frac{1}{2}H$ and they satisfy the initial conditions. Finally, unwinding the definitions of $\tilde{\beta}(t)$ and $\tilde{\Sigma}(t)$ gives the stated result. $\square$

### 5 Calculations for local information

#### 5.1 Master convolution equations

Collecting all parameters in one vector $\vec{\beta}$, with components indexed by $k$ and $l$, handles parameters shared across transitions (such as a single background mortality rate applying to every living state) without further bookkeeping. We denote by $\pi_s(t;\vec{\beta})$ the arrival time distributions of individuals to state $s$ for a value $\vec{\beta}$ of the model parameters. As shown in Proposition 1, these distributions satisfy a system of flow conservation equations with a source term coming from the initial state $s_0$ at time 0:

$$\pi(t;\vec{\beta}) - (f * \pi)(t;\vec{\beta}) = \delta_0(t) \cdot e_{s_0},$$

where $\mathbf{f}$ is the matrix of sub-densities with entries $\mathbf{f}_{sq}(t;\vec{\beta}) = f_{qs}(t;\vec{\beta})$, whose Laplace transform is the matrix $F(\omega;\vec{\beta})$ of Corollary 2 in the main text, $\delta_0(t)$ is the Dirac delta distribution concentrated at $t = 0$ and $e_{s_0}$ is the canonical vector of the initial state (i.e., the vector with value one in its $s_0$-th component and zero in the rest). From Corollary 3, the population profile functions are obtained from the arrival time distributions by means of convolution with the corresponding holding time distributions:

$$p_s(t;\vec{\beta}) = (\pi_s(\cdot;\vec{\beta}) * S_s(\cdot;\vec{\beta}))(t).$$

If we denote $\Pi_s^{(0)}(\omega;\vec{\beta}) := \mathcal{L}_{\pi_s(\cdot;\vec{\beta})}(\omega)$, the corresponding equations in the frequency domain are:

$$\begin{aligned} &[\mathrm{Id} - F(\omega;\vec{\beta})]\Pi^{(0)}(\omega;\vec{\beta}) = e_{s_0} \\ \Longrightarrow\ &\Pi^{(0)}(\omega;\vec{\beta})_s = [A(\omega;\vec{\beta})^{-1} \cdot e_{s_0}]_s, \end{aligned}$$

$$\mathcal{L}_{p_s}(\omega;\vec{\beta}) = \Pi_s^{(0)}(\omega;\vec{\beta}) \cdot \mathcal{L}_{S_s}(\omega;\vec{\beta}),$$

where $A(\omega;\vec{\beta}) := \mathrm{Id} - F(\omega;\vec{\beta})$. In the following calculations, we will also denote by $\Pi^{(d)}$ the derivatives of order $d$ for these functions with respect to their $\vec{\beta}$ parameters.

The relationship between hazards $h$ and density functions $f$ is given by

$$f_{qr}(t;\vec{\beta}) = h_{qr}(t;\vec{\beta}) \cdot e^{-H_q(t;\vec{\beta})},$$

where, as before, $H_q(t;\vec{\beta}) = \int_0^t \sum_r h_{qr}(u;\vec{\beta})\, du$ is the cumulative hazard of state $q$ at time $t$, and $S_q(t;\vec{\beta}) = e^{-H_q(t;\vec{\beta})}$.

#### 5.2 Gradients

Starting from the previous calculations, we take a particular parameter component $k$ and differentiate on $\beta_k$:

$$\begin{aligned} A(\omega;\vec{\beta})\Pi^{(0)}(\omega;\vec{\beta}) &= e_{s_0} \\ \Longrightarrow\ \frac{\partial A}{\partial \beta_k}(\omega;\vec{\beta})\Pi^{(0)}(\omega;\vec{\beta}) + A(\omega;\vec{\beta})\Pi_{k\cdot}^{(1)}(\omega;\vec{\beta}) &= 0 \\ \Longrightarrow\ [A(\omega;\vec{\beta})^{-1} \cdot \mathcal{L}_{\frac{\partial f}{\partial \beta_k}}(\omega;\vec{\beta}) \cdot \Pi^{(0)}(\omega;\vec{\beta})]_s &= \Pi_{ks}^{(1)}(\omega;\vec{\beta}), \end{aligned}$$

where we have used

$$A(\omega;\vec{\beta}) = \mathrm{Id} - \mathcal{L}_f(\omega;\vec{\beta}) \implies \frac{\partial A}{\partial \beta_k}(\omega;\vec{\beta}) = -\frac{\partial \mathcal{L}_f}{\partial \beta_k}(\omega;\vec{\beta}) = -\mathcal{L}_{\frac{\partial f}{\partial \beta_k}}(\omega;\vec{\beta})$$

by differentiation under the integral sign in the Laplace transform definition. The $f$-quantities on display can also be expressed in terms of the hazard rate functions:

$$\frac{\partial f_{qr}}{\partial \beta_k}(t;\vec{\beta}) = \left[\frac{\partial h_{qr}}{\partial \beta_k}(t;\vec{\beta}) - h_{qr}(t;\vec{\beta}) \cdot \frac{\partial H_q}{\partial \beta_k}(t;\vec{\beta})\right] \cdot e^{-H_q(t;\vec{\beta})},$$

and the gradients of the population profile functions:

$$\frac{\partial \mathcal{L}_{p_s}}{\partial \beta_k}(\omega;\vec{\beta}) = \Pi^{(1)}_{ks}(\omega;\vec{\beta}) \cdot \mathcal{L}_{S_s}(\omega;\vec{\beta}) + \Pi^{(0)}_{s}(\omega;\vec{\beta}) \cdot \frac{\partial \mathcal{L}_{S_s}}{\partial \beta_k}(\omega;\vec{\beta}).$$

##### 5.3 Hessians

Yet again, we take a parameter component $l$ and differentiate on $\beta_l$ (we omit the function arguments $(\omega;\vec{\beta})$ for clarity) and replace the expressions obtained for the first-order derivatives of $A$ in the previous section.

$$\begin{aligned}
\frac{\partial A}{\partial \beta_k}\Pi^{(0)} + A\Pi^{(1)}_{k\cdot} &= 0 \\
\implies \frac{\partial^2 A}{\partial \beta_l \partial \beta_k}\Pi^{(0)} + \frac{\partial A}{\partial \beta_k}\Pi^{(1)}_{l\cdot} + \frac{\partial A}{\partial \beta_l}\Pi^{(1)}_{k\cdot} + A\Pi^{(2)}_{lk\cdot} &= 0 \\
\implies \left[A^{-1} \cdot \left(\mathcal{L}_{\frac{\partial^2 f}{\partial \beta_l \partial \beta_k}} \cdot \Pi^{(0)} + \mathcal{L}_{\frac{\partial f}{\partial \beta_k}} \cdot \Pi^{(1)}_{l\cdot} + \mathcal{L}_{\frac{\partial f}{\partial \beta_l}} \cdot \Pi^{(1)}_{k\cdot}\right)\right]_s &= \Pi^{(2)}_{lks}.
\end{aligned}$$

Similarly as before, we can also express the $f$-quantities in terms of the hazard rate functions:

$$\begin{aligned}
\frac{\partial^2 f_{qr}}{\partial \beta_l \partial \beta_k}(t) &= \left[\left(\frac{\partial^2 h_{qr}}{\partial \beta_l \partial \beta_k} - \frac{\partial h_{qr}}{\partial \beta_l} \cdot \frac{\partial H_q}{\partial \beta_k} - h_{qr} \cdot \frac{\partial^2 H_q}{\partial \beta_l \partial \beta_k}\right) - \left(\frac{\partial h_{qr}}{\partial \beta_k} - h_{qr} \cdot \frac{\partial H_q}{\partial \beta_k}\right) \cdot \frac{\partial H_q}{\partial \beta_l}\right] \cdot e^{-H_q(t)} \\
&= \left[\left(\frac{\partial^2 h_{qr}}{\partial \beta_l \partial \beta_k} - h_{qr} \cdot \frac{\partial^2 H_q}{\partial \beta_l \partial \beta_k}\right) - \left(\frac{\partial h_{qr}}{\partial \beta_l} \cdot \frac{\partial H_q}{\partial \beta_k} + \frac{\partial h_{qr}}{\partial \beta_k} \cdot \frac{\partial H_q}{\partial \beta_l}\right) + h_{qr} \cdot \frac{\partial H_q}{\partial \beta_l} \cdot \frac{\partial H_q}{\partial \beta_k}\right] \cdot e^{-H_q(t)},
\end{aligned}$$

and the Hessians of the population profile functions:

$$\frac{\partial^2 \mathcal{L}_{p_s}}{\partial \beta_l \partial \beta_k} = \Pi^{(2)}_{lks} \cdot \mathcal{L}_{S_s} + \Pi^{(1)}_{ks} \cdot \frac{\partial \mathcal{L}_{S_s}}{\partial \beta_l} + \Pi^{(1)}_{ls} \cdot \frac{\partial \mathcal{L}_{S_s}}{\partial \beta_k} + \Pi^{(0)}_{s} \cdot \frac{\partial \mathcal{L}_{S_s}}{\partial \beta_l \partial \beta_k}.$$

#### 6 SEER data

We used the SEER*Stat Database [2] to construct the incidence and mortality data for our calibration experiment. We considered cases of patients diagnosed with urinary bladder cancer, observed between 2004 and 2021, and classified the diagnosis stages as follows:

- Early: In situ/Localized,

- Medium: Regional,
- Late: Distant.

For simplicity in the calculations, we ignored possible cohort effects and used the formulas described in [3] to calculate the crude rates and standard errors associated with the observed clinical incidence, as well as preclinical and clinical mortality, for each of the stages of the model. In the calculation of preclinical mortality rates and confidence intervals, we use a cutoff of death occurring at most 2 months after diagnosis. The incidence and preclinical (deaths-at-diagnosis) targets use the bladder cancers diagnosed in 2004 to 2021 in the SEER 17 registries (Incidence – SEER Research Data, 17 Registries, Nov 2023 Sub), and the clinical mortality targets use the deaths of the same registries' cases in the matching incidence-based mortality database. All targets share one denominator: the person-years of the SEER 17 registries in 2004 to 2021, by single year of age. Combined Summary Stage is recorded only for cases diagnosed from 2004, so the deaths of cases diagnosed in 2000 to 2003, about 20% of the bladder cancer deaths, have no stage and enter no target, as do the 6% of deaths with unknown stage. The clinical mortality targets therefore omit deaths among cases diagnosed before 2004 and are biased downward, most in the first years of the period.

Tables S1 and S2 show the resulting incidence and mortality rates and their standard errors.

##### 6.1 Background mortality

In the application, we take the background survival from the observed survival of the US population, $S_0(t) \approx S_{\mathrm{US}}(t)$, using the 2021 US life table, and set the model survival $\mathcal{S}(t)$ to one in the weights of Section 2.5.4 of the main text and in the other-cause mortality rate $\lambda_s^{(\mathrm{ext})}$. Strictly, total survival should equal $S_{\mathrm{US}}$, which would require $S_0 = S_{\mathrm{US}}/\mathcal{S}$, so that deaths from the modeled cancer are not counted twice; the approximation neglects the cancer's own share of all-cause mortality. In our estimates $\mathcal{S}(t)$ stays above 0.996 through age 80, so the approximation does not change any target by more than 0.5%. Because $S_{\mathrm{US}}$ is data, the background term does not depend on the model parameters.

The life table gives $S_{\mathrm{US}}$ at integer ages. We treat it as constant on the half-year on either side of each age, and evaluate the weighted interval integrals of Section 2.5.4 of the main text exactly for that step function, from the cumulative arrivals at the integer and half-year points.

#### 7 Verification and benchmark details

##### 7.1 Independent reference implementation

Every post-onset transition of the bladder cancer model has a constant hazard, so the process restricted to the post-onset states $\{U_1, U_2, U_3, T_1, T_2, T_3, D_c\}$ is a time-homogeneous Markov chain with generator $Q$, and the vector of occupancies $p(t)$ of these states solves

$$p'(t) = Q^{\top} p(t) + f_{HU_1}(t)\, e_{U_1}, \qquad p(0) = 0,$$

where $f_{HU_1}$ is the Weibull onset density. We solved this system with the exponential midpoint scheme $p(t+h) = e^{Q^{\top} h} p(t) + h\, f_{HU_1}(t + h/2)\, e^{Q^{\top} h/2} e_{U_1}$ with $h = 1/256$ years, and we assembled the interval targets of Section 2.5 of the main text by trapezoidal integration on the same grid. Halving the step to $h = 1/512$ changed no target by more than a relative $5.9 \times 10^{-6}$. The implementation is written in base R, shares no code with SMORFS, and is included in the software deposit.

##### 7.2 Comparators

The cohort approximation uses a cycle of length $\Delta$ (one month or one year), converts each rate to a per-cycle probability $1 - e^{-\text{rate}\cdot\Delta}$ with destinations split in proportion to the competing rates, takes the per-cycle onset probability from the Weibull cumulative hazard, and uses mid-cycle survivors as the denominator. The microsimulation draws onset ages for $2 \times 10^6$ individuals from the Weibull distribution and post-onset histories by competing exponential draws, and computes person-years exactly from each individual's age at death. All comparisons use the parameter values of Table 2 of the original analysis, with background mortality removed.

##### 7.3 Results

Figure S1 shows the maximum relative error of the SMORFS targets against the reference as the number of series terms grows. The 120 terms are 24 half-periods of $\ell = 5$ terms, with the acceleration applied to the half-period partial sums. From 100 terms on it stays at $7.9 \times 10^{-6}$, the reference's own accuracy ($5.9 \times 10^{-6}$ by step halving) rather than the inversion's; with 60 terms it would be $7.0 \times 10^{-5}$. At 120 terms, the pointwise relative error of the arrival densities at ages 35 to 80 is $1.7 \times 10^{-8}$. Near $t = 0$ the inversion error alternates in sign at about $10^{-12}$ in absolute terms, while the true densities are between $10^{-26}$ and $4 \times 10^{-11}$, so relative error is not meaningful there; no target lies in this region.

With background mortality included as in estimation, the analytic gradient agrees with central finite differences of the objective (step $10^{-4}$ on the log scale) to a maximum relative error of $8.6 \times 10^{-8}$ over all 13 parameters, at a point displaced by 0.1 on the log scale from the maximum likelihood estimate. At the estimate, the analytic Hessian agrees with central differences of an accurate gradient to a relative error of $9.7 \times 10^{-7}$ in the Frobenius norm, and it is positive definite, with smallest eigenvalue 60.5.

All timings in Table 3 of the main text are single-threaded medians of 10 runs on an Apple M4 Max (12 performance and 4 efficiency cores, 128 GB) running macOS 26.6.2, with Python 3.12.15, NumPy 2.4.4, SciPy 1.17.1, R 4.5.3, and the C++ library compiled with Apple clang 21.0.0 and linked to Apple's Accelerate framework.

#### 8 Software and example model specification

SMORFS creates states and transitions from parametric families; computes arrival densities, cumulative distributions, and population profile functions with their first and second derivatives; applies gradient, Newton, and saddle-free Newton steps; assesses local identifiability from first- and second-order information; and performs Bayesian estimation by LIMIS. All frequency-domain calculations are hidden from the user. Listing 1 shows the specification of the bladder cancer model of the main text.

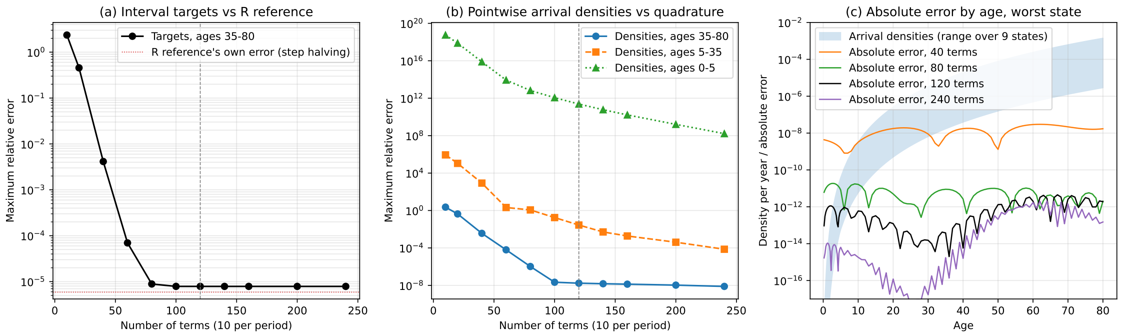


Figure S1: Maximum relative error of the SMORFS targets against the independent reference, by number of series terms (log scale). The horizontal line marks the reference's own accuracy.

| | Clinical incidence | | | Clinical mortality | | |
|---|---|---|---|---|---|---|
| | Early | Medium | Late | Early | Medium | Late |
| 35 years | 0.912 (0.0664) | 0.0821 (0.0199) | 0.0579 (0.0167) | 0.0241 (0.0108) | * | * |
| 36 years | 1.01 (0.0698) | 0.0629 (0.0174) | 0.0581 (0.0168) | 0.0339 (0.0128) | 0.0435 (0.0145) | 0.0339 (0.0128) |
| 37 years | 1.26 (0.0783) | 0.0923 (0.0212) | 0.0486 (0.0154) | 0.0243 (0.0109) | 0.0243 (0.0109) | 0.0291 (0.0119) |
| 38 years | 1.27 (0.0788) | 0.102 (0.0223) | 0.078 (0.0195) | 0.0341 (0.0129) | 0.0488 (0.0154) | 0.0488 (0.0154) |
| 39 years | 1.48 (0.0848) | 0.0975 (0.0218) | 0.0828 (0.0201) | 0.039 (0.0138) | 0.0487 (0.0154) | 0.039 (0.0138) |
| 40 years | 1.8 (0.0938) | 0.127 (0.025) | 0.132 (0.0254) | 0.0686 (0.0183) | 0.0294 (0.012) | 0.0539 (0.0162) |
| 41 years | 2.05 (0.0999) | 0.18 (0.0296) | 0.122 (0.0243) | 0.0633 (0.0176) | 0.0438 (0.0146) | 0.0925 (0.0212) |
| 42 years | 2.11 (0.101) | 0.214 (0.0323) | 0.185 (0.03) | 0.0827 (0.02) | 0.0924 (0.0212) | 0.0778 (0.0195) |
| 43 years | 2.74 (0.115) | 0.175 (0.0291) | 0.209 (0.0318) | 0.0971 (0.0217) | 0.0923 (0.0212) | 0.141 (0.0261) |
| 44 years | 3.31 (0.127) | 0.291 (0.0375) | 0.155 (0.0274) | 0.131 (0.0252) | 0.0872 (0.0206) | 0.15 (0.027) |
| 45 years | 3.76 (0.135) | 0.364 (0.0421) | 0.228 (0.0333) | 0.16 (0.0279) | 0.102 (0.0223) | 0.17 (0.0287) |
| 46 years | 4.34 (0.145) | 0.41 (0.0445) | 0.256 (0.0351) | 0.125 (0.0246) | 0.164 (0.0281) | 0.188 (0.0301) |
| 47 years | 4.8 (0.152) | 0.495 (0.0488) | 0.346 (0.0408) | 0.231 (0.0333) | 0.202 (0.0311) | 0.207 (0.0315) |
| 48 years | 5.74 (0.166) | 0.58 (0.0528) | 0.413 (0.0445) | 0.211 (0.0318) | 0.24 (0.0339) | 0.278 (0.0365) |
| 49 years | 6.57 (0.177) | 0.653 (0.0558) | 0.429 (0.0452) | 0.238 (0.0337) | 0.224 (0.0327) | 0.372 (0.0421) |
| 50 years | 7.75 (0.194) | 0.765 (0.0609) | 0.47 (0.0477) | 0.32 (0.0393) | 0.32 (0.0393) | 0.315 (0.039) |
| 51 years | 8.86 (0.207) | 0.747 (0.0602) | 0.587 (0.0534) | 0.383 (0.0431) | 0.277 (0.0366) | 0.383 (0.0431) |
| 52 years | 9.96 (0.222) | 1.01 (0.0705) | 0.561 (0.0526) | 0.379 (0.0432) | 0.34 (0.0409) | 0.438 (0.0465) |
| 53 years | 12 (0.244) | 1.1 (0.0737) | 0.749 (0.0609) | 0.575 (0.0534) | 0.436 (0.0465) | 0.486 (0.0491) |
| 54 years | 13.3 (0.258) | 1.28 (0.0802) | 0.912 (0.0676) | 0.616 (0.0556) | 0.586 (0.0542) | 0.666 (0.0578) |
| 55 years | 15.3 (0.279) | 1.43 (0.0855) | 0.903 (0.0679) | 0.689 (0.0593) | 0.582 (0.0545) | 0.694 (0.0595) |
| 56 years | 16.9 (0.295) | 1.78 (0.0959) | 1.17 (0.0776) | 0.919 (0.0689) | 0.712 (0.0606) | 0.78 (0.0634) |
| 57 years | 19.9 (0.323) | 1.84 (0.0983) | 1.25 (0.081) | 1.1 (0.0759) | 0.797 (0.0646) | 0.734 (0.062) |
| 58 years | 22 (0.344) | 2.19 (0.108) | 1.28 (0.0831) | 1.15 (0.0788) | 1.03 (0.0746) | 1.1 (0.0769) |
| 59 years | 25.2 (0.375) | 2.24 (0.112) | 1.65 (0.096) | 1.53 (0.0924) | 1.11 (0.0785) | 1.22 (0.0824) |
| 60 years | 28.3 (0.406) | 2.5 (0.121) | 1.85 (0.104) | 1.81 (0.103) | 1.04 (0.0777) | 1.27 (0.086) |
| 61 years | 31.6 (0.435) | 2.96 (0.133) | 2.03 (0.11) | 1.95 (0.108) | 1.13 (0.0822) | 1.37 (0.0904) |
| 62 years | 35.8 (0.472) | 3.28 (0.143) | 1.96 (0.11) | 2.39 (0.122) | 1.46 (0.0952) | 1.52 (0.0972) |
| 63 years | 39.8 (0.509) | 3.38 (0.148) | 2.23 (0.121) | 2.73 (0.133) | 1.62 (0.103) | 1.73 (0.106) |
| 64 years | 42.5 (0.539) | 3.55 (0.156) | 2.61 (0.134) | 3.11 (0.146) | 1.85 (0.112) | 1.67 (0.107) |
| 65 years | 50.9 (0.604) | 4.43 (0.178) | 2.88 (0.144) | 3.72 (0.163) | 1.99 (0.119) | 2.17 (0.125) |
| 66 years | 55.2 (0.645) | 4.63 (0.187) | 3.01 (0.151) | 4.1 (0.176) | 2.06 (0.124) | 2.39 (0.134) |
| 67 years | 58.7 (0.682) | 4.84 (0.196) | 3.14 (0.158) | 4.77 (0.194) | 2.16 (0.131) | 2.06 (0.128) |
| 68 years | 63.5 (0.727) | 5.56 (0.215) | 3.24 (0.164) | 5.37 (0.211) | 2.28 (0.138) | 2.08 (0.132) |
| 69 years | 68.9 (0.781) | 5.57 (0.222) | 3.61 (0.179) | 6.33 (0.237) | 2.6 (0.152) | 2.74 (0.156) |
| 70 years | 75.3 (0.841) | 6.3 (0.243) | 4.26 (0.2) | 7.34 (0.263) | 2.61 (0.157) | 2.76 (0.161) |
| 71 years | 80.4 (0.892) | 6.98 (0.263) | 4.34 (0.207) | 8.44 (0.289) | 3.5 (0.186) | 2.82 (0.167) |
| 72 years | 87.9 (0.96) | 7 (0.271) | 4.7 (0.222) | 8.27 (0.295) | 3.42 (0.189) | 3.29 (0.186) |
| 73 years | 91.7 (1) | 7.35 (0.284) | 5 (0.234) | 9.88 (0.329) | 3.33 (0.191) | 3.82 (0.205) |
| 74 years | 101 (1.1) | 8.66 (0.322) | 5.39 (0.254) | 11.9 (0.377) | 4.16 (0.223) | 3.87 (0.215) |
| 75 years | 109 (1.18) | 9.15 (0.343) | 6.03 (0.278) | 12.4 (0.398) | 4.52 (0.241) | 4.14 (0.231) |
| 76 years | 115 (1.25) | 9.98 (0.367) | 7.1 (0.309) | 15.2 (0.453) | 4.42 (0.244) | 4.9 (0.257) |
| 77 years | 118 (1.29) | 9.84 (0.374) | 6.25 (0.298) | 16.8 (0.488) | 5.38 (0.276) | 5.08 (0.269) |
| 78 years | 126 (1.38) | 9.97 (0.388) | 6.41 (0.311) | 19.1 (0.537) | 5.13 (0.278) | 5.04 (0.276) |
| 79 years | 131 (1.45) | 10.4 (0.41) | 7.76 (0.353) | 21.4 (0.588) | 5.23 (0.29) | 5.62 (0.301) |
| 80 years | 133 (1.52) | 11.1 (0.438) | 7.46 (0.358) | 23.8 (0.641) | 6.34 (0.33) | 5.22 (0.3) |

Table S1: SEER clinical incidence and mortality rates, with corresponding standard errors. Cells marked * are based on 1 to 4 events and are suppressed, as the SEER data-use agreement requires; they are used in the calibration. Cells marked - have no events.

| | Preclinical mortality | | | Preclinical other-cause mortality | | |
|---|---|---|---|---|---|---|
| | Early | Medium | Late | Early | Medium | Late |
| 35 years | * | - | * | - | - | - |
| 36 years | - | - | * | - | - | - |
| 37 years | - | - | - | * | - | * |
| 38 years | - | - | * | * | - | - |
| 39 years | * | - | * | - | * | - |
| 40 years | * | * | * | - | - | - |
| 41 years | * | * | 0.0292 (0.0119) | * | - | - |
| 42 years | * | * | 0.0389 (0.0138) | * | - | - |
| 43 years | * | * | 0.0291 (0.0119) | * | - | * |
| 44 years | 0.0242 (0.0108) | * | 0.0291 (0.0119) | * | - | * |
| 45 years | * | * | 0.0291 (0.0119) | * | - | - |
| 46 years | * | * | 0.0531 (0.016) | * | - | * |
| 47 years | * | * | 0.0625 (0.0173) | * | - | - |
| 48 years | 0.024 (0.0107) | 0.0288 (0.0117) | 0.0624 (0.0173) | - | - | * |
| 49 years | * | * | 0.0858 (0.0202) | 0.0238 (0.0107) | - | * |
| 50 years | 0.0339 (0.0128) | * | 0.0871 (0.0205) | * | * | * |
| 51 years | 0.0485 (0.0153) | 0.0243 (0.0109) | 0.102 (0.0222) | 0.0291 (0.0119) | - | 0.0243 (0.0109) |
| 52 years | * | 0.0246 (0.011) | 0.123 (0.0246) | 0.0492 (0.0156) | - | * |
| 53 years | 0.0595 (0.0172) | 0.0248 (0.0111) | 0.129 (0.0253) | 0.0546 (0.0164) | - | * |
| 54 years | 0.0852 (0.0207) | 0.0251 (0.0112) | 0.21 (0.0325) | 0.0601 (0.0174) | * | * |
| 55 years | 0.051 (0.0161) | 0.0561 (0.0169) | 0.214 (0.0331) | 0.0714 (0.0191) | * | * |
| 56 years | 0.0465 (0.0155) | 0.031 (0.0126) | 0.268 (0.0372) | 0.0361 (0.0137) | * | 0.0258 (0.0115) |
| 57 years | 0.0786 (0.0203) | 0.0681 (0.0189) | 0.283 (0.0385) | 0.0786 (0.0203) | * | * |
| 58 years | 0.0808 (0.0209) | 0.0646 (0.0187) | 0.258 (0.0373) | 0.108 (0.0241) | 0.0377 (0.0142) | * |
| 59 years | 0.106 (0.0243) | 0.0502 (0.0167) | 0.368 (0.0453) | 0.112 (0.025) | * | 0.0558 (0.0176) |
| 60 years | 0.175 (0.0319) | 0.0583 (0.0184) | 0.437 (0.0504) | 0.146 (0.0291) | * | 0.0291 (0.013) |
| 61 years | 0.185 (0.0333) | 0.0657 (0.0198) | 0.341 (0.0451) | 0.197 (0.0343) | * | * |
| 62 years | 0.149 (0.0304) | 0.0683 (0.0206) | 0.416 (0.0508) | 0.217 (0.0368) | * | 0.0559 (0.0186) |
| 63 years | 0.183 (0.0345) | 0.157 (0.0319) | 0.515 (0.058) | 0.209 (0.0369) | * | * |
| 64 years | 0.253 (0.0415) | 0.102 (0.0264) | 0.56 (0.0618) | 0.3 (0.0453) | 0.041 (0.0167) | 0.0819 (0.0236) |
| 65 years | 0.38 (0.0522) | 0.136 (0.0313) | 0.653 (0.0684) | 0.395 (0.0532) | 0.043 (0.0176) | 0.0789 (0.0238) |
| 66 years | 0.324 (0.0494) | 0.158 (0.0345) | 0.7 (0.0726) | 0.452 (0.0583) | 0.0452 (0.0184) | 0.0979 (0.0271) |
| 67 years | 0.412 (0.0571) | 0.111 (0.0296) | 0.72 (0.0755) | 0.546 (0.0658) | 0.0475 (0.0194) | 0.15 (0.0345) |
| 68 years | 0.349 (0.0539) | 0.216 (0.0424) | 0.691 (0.0758) | 0.449 (0.0611) | 0.0915 (0.0276) | 0.116 (0.0311) |
| 69 years | 0.549 (0.0698) | 0.257 (0.0477) | 0.992 (0.0938) | 0.585 (0.072) | 0.0975 (0.0294) | 0.221 (0.0443) |
| 70 years | 0.564 (0.0728) | 0.207 (0.0441) | 1.09 (0.101) | 0.705 (0.0814) | 0.122 (0.0339) | 0.273 (0.0506) |
| 71 years | 0.584 (0.0761) | 0.238 (0.0485) | 1.08 (0.103) | 0.743 (0.0858) | 0.119 (0.0343) | 0.129 (0.0357) |
| 72 years | 0.766 (0.0896) | 0.294 (0.0555) | 1.1 (0.107) | 0.944 (0.0995) | 0.105 (0.0332) | 0.21 (0.0469) |
| 73 years | 0.888 (0.0987) | 0.318 (0.0591) | 1.43 (0.125) | 1.09 (0.109) | 0.0987 (0.0329) | 0.154 (0.041) |
| 74 years | 1.02 (0.11) | 0.538 (0.0801) | 1.35 (0.127) | 1.24 (0.122) | 0.167 (0.0447) | 0.191 (0.0478) |
| 75 years | 1.11 (0.119) | 0.514 (0.0813) | 1.77 (0.151) | 1.18 (0.123) | 0.219 (0.053) | 0.18 (0.0481) |
| 76 years | 1.51 (0.143) | 0.606 (0.0904) | 2.1 (0.168) | 1.55 (0.144) | 0.323 (0.066) | 0.337 (0.0674) |
| 77 years | 1.49 (0.146) | 0.497 (0.084) | 2.07 (0.172) | 1.68 (0.154) | 0.199 (0.0531) | 0.327 (0.0681) |
| 78 years | 2.25 (0.184) | 0.769 (0.108) | 1.96 (0.172) | 1.81 (0.165) | 0.272 (0.064) | 0.392 (0.0769) |
| 79 years | 2.19 (0.188) | 0.757 (0.11) | 2.48 (0.2) | 2.53 (0.202) | 0.258 (0.0644) | 0.37 (0.0772) |
| 80 years | 2.95 (0.225) | 0.982 (0.13) | 2.24 (0.196) | 2.48 (0.207) | 0.379 (0.0808) | 0.5 (0.0928) |

Table S2: SEER preclinical mortality rates, from the cancer and from other causes, with corresponding standard errors. Cells marked * are based on 1 to 4 events and are suppressed, as the SEER data-use agreement requires; they are used in the calibration. Cells marked - have no events.

```
# Inherit from the base semi-Markov model class
class BladderModel(SemiMarkov):
    def __init__(self, name):
        # Call the base class constructor to pre-initialize data structures
        super().__init__(name)

        # Create states
        self.H = State("H")
        self.U1 = State("U1")
        self.U2 = State("U2")
        [...]

        # Add states to the model
        for s in [self.H, self.U1, self.U2, self.U3, self.T1, self.T2, self.T3, self.Dc]:
            self.add_state(s)

        # Set initial and terminal states
        self.set_initial(self.H)
        self.Dc.terminal = True

        # Create transitions with their names and initial parameter values
        lambda0 = 1e-6
        self.R_inc = WeibullStable("Preclinical onset", k=shape, s=scale)
        self.R_U1_U2 = ExponentialStable("U1->U2", rate=6e2*lambda0)
        self.R_U1_T1 = ExponentialStable("U1->T1", rate=1e3*lambda0)
        self.R_U2_U3 = ExponentialStable("U2->U3", rate=3e5*lambda0)
        self.R_U2_T2 = ExponentialStable("U2->T2", rate=3e5*lambda0)
        [...]

        # Add transitions from corresponding states
        self.H.add_transition(self.U1, self.R_inc)
        self.U1.add_transition(self.U2, self.R_U1_U2)
        self.U1.add_transition(self.T1, self.R_U1_T1)
        [...]

        # Automatically calculate necessary data from the final model structure
        self.update()
```

Listing 1: Specification code for the urinary bladder cancer model.
